# Beyond willingness to donate: a cross-sectional online survey to understand urgent blood-donor mobilisability among digitally reachable adults in Ghana

**DOI:** 10.64898/2026.08.27.26361538

**Authors:** Honghui Shen, Isaiah Awintuen Agorinya, Martin Amogre Ayanore, Markus Brede, Adriane Chapman, Michael Head

## Abstract

**Objectives:** To identify factors associated with anticipated engagement in urgent blood-donor mobilisation among digitally reachable adults in Ghana.

**Background:** Digital tools may facilitate contact with potential blood donors during emergencies and blood shortages, but digital reachability does not ensure engagement with urgent requests.

**Methods:** We conducted a cross-sectional online survey across Ghana’s 16 regions (September 2025–January 2026), recruiting participants via Facebook advertising and snowball sampling. Factors associated with urgent blood-donor mobilisability were assessed under four criteria: future-donation willingness; willingness to install a trusted donation app; willingness to respond to a trusted urgent-request; and practical flexibility to leave current activities. Descriptive analyses and multivariable logistic regression examined prevalence and associated factors.

**Results:** Of 1,067 respondents, 577 (54.1%) met all four criteria, compared with 91.8% reporting high future-donation willingness. Trusted-app installation willingness (83.2%) and trusted-request response willingness (82.7%) exceeded practical flexibility (66.6%). High mobilisability was associated with formal health-system trust (AOR 3.95, 95% CI 2.08–7.50), digital-response readiness (AOR 2.26, 1.66–3.08), donation knowledge (AOR 1.42, 1.03–1.97) and previous donation (AOR 1.47, 1.08–2.01). Women (AOR 0.60, 0.43–0.83), those with work-schedule barriers (AOR 0.43, 0.29–0.66) and those reporting >30-minute night-time travel to the nearest healthcare facility (AOR 0.66, 0.45–0.96) had lower adjusted odds.

**Conclusions:** Digital reachability and stated donation willingness may overestimate the population pool available for emergency donation. Digital blood-donor solutions should consider verifiable health-system requests, account for digital-response readiness and current availability, and connect willing individuals with accessible collection options and transport support where needed.

## INTRODUCTION

Safe and timely access to blood remains a persistent challenge for global health systems, particularly in low- and middle-income countries (LMICs) [1,2]. Blood transfusion is essential for managing obstetric haemorrhage, severe anaemia including after infections such as malaria, trauma, surgery, sickle-cell disease and other acute and chronic conditions, particularly relevant in sub-Saharan Africa and other LMICs.

However, recent World Health Organization (WHO) data indicate substantial global inequalities in blood availability. Of approximately 120 million blood donations collected worldwide annually, 36% are collected in HICs, which account for only 15% of the global population. Median donation rates also vary markedly by income level, from 28.9 donations per 1,000 population in HICs to 8.5 in lower-middle-income countries and 4.5 in low-income countries [1]. A 2019 global modelling study estimated that 119 of 195 countries did not have sufficient blood supply to meet clinical need, with unmet need concentrated in LMICs. It also suggested that all countries in central, eastern and western sub-Saharan Africa (SSA) had insufficient blood to meet estimated transfusion needs [3].

These gaps are especially consequential in emergency-care contexts, where the need for blood can arise suddenly and where delays may directly affect survival. Postpartum haemorrhage (PPH) is a major example to illustrate the time-critical nature of this problem. Globally, an estimated 27 million women experience PPH each year, with around 43000 deaths [4]. Sub-Saharan Africa accounted for around 70% of global maternal deaths in 2023, with haemorrhage accounting for an estimated 28% of maternal deaths in the region [5,6]. Most haemorrhage-related maternal deaths occur soon after childbirth, and deaths from obstetric bleeding are largely preventable with timely diagnosis, effective first-line treatment, escalation of care, and timely access to safe blood products [7,8]. Evidence from the SSA region suggests that ineffective blood transfusion services make a substantial contribution to maternal haemorrhage deaths, attributing at least one-quarter of such preventable deaths to lack of rapid access to blood [9,10]. Urgent blood need is therefore a recurrent challenge within ordinary hospital practice [2,11], and is not confined to rural settings. Across sub-Saharan Africa, shortages and delays in emergency transfusion have been documented even in major urban referral hospitals, while evidence from Ghana similarly indicates unmet transfusion need in urban tertiary care alongside substantial emergency transfusion requirements in regional settings [12–15].

A growing body of evidence from sub-Saharan Africa shows that blood-supply constraints are shaped by both service-level and donor-level factors. Reported challenges include low recruitment and retention of voluntary donors, reliance on family replacement donation, donor ineligibility related to anaemia and transfusion-transmissible infections, limited blood-service resources, and insufficient public communication [16–18]. In Ghana, West Africa, studies report similar patterns, suggesting that willingness to donate is influenced by knowledge, motivation, fear, previous donation experience, perceived health risks, and practical barriers [19,20].

Digital coordination may offer one route to strengthening donor mobilisation, particularly where mobile connectivity is now widespread. WHO’s global digital health strategy emphasises the potential of digital technologies to support health-system strengthening [21]. For blood donation, digital tools could potentially support faster identification of eligible donors, more targeted alerts, two-way communication, location-aware coordination, and real-time updates between hospitals and potential donors [22,23].

Ghana has substantial digital communication infrastructure. By December 2025, the National Communications Authority reported 28.7 million active mobile data subscriptions, representing a penetration rate of 85.0%, alongside 26.9 million connected smartphones [24]. This creates a promising opportunity to use digital tools to expand the reach for blood donations.

Nevertheless, digital reach alone does not guarantee digital engagement. A person may own a smartphone, use mobile internet, and be reachable through a digital platform, yet still be unwilling to use a blood-donation app, distrust a digital request, or be unable or unlikely to respond promptly when an urgent request for blood is sent. In urgent blood-donor mobilisation, therefore, the key question is not only whether people are digitally reachable, but whether they can plausibly be mobilised.

Existing research provides limited evidence for this question. Mobile blood-donation studies have described how apps can support donor identification, recruitment, retention, scheduling, reminders, geolocation, gamification and communication between donors and blood services [22,23,25]. In South Africa, survey research has examined blood donors’ app usage behaviour and perceptions of a proposed blood donation app, suggesting that app-based engagement may support donor recruitment and retention [26]. Other digital studies have examined social-media recruitment, algorithmic donor matching, online donor communities, chatbots and behaviour-change support systems [27–30]. However, much of the evidence focuses on routine donation, donor retention, or general willingness to use a blood-donation app. Less is known about digitally coordinated urgent blood-donor mobilisation, particularly in African health-system settings where local blood stocks may often be insufficient and donors may be sought in response to a time-critical emergency.

Existing studies from Ghana and other sub-Saharan African settings have identified established influences on blood donation, including knowledge, motivation, fear, previous donation experience and practical barriers. However, these factors have rarely been examined alongside the digital and operational conditions required for urgent mobilisation, such as mobile-phone use patterns, notification-checking and response behaviour, app-installation confidence, trust in digital-request sources, and the practical ability to leave current activities quickly after receiving an urgent request [16,18,19,26]. This leaves an important evidence gap: we know relatively little about who is likely to engage with urgent digital blood-donor requests, and what individual, social, trust-related, and practical factors may shape this engagement. This study addresses this gap using survey data from digitally reachable adults across Ghana. We examine factors associated with potential engagement with digitally coordinated urgent blood-donor mobilisation. Our focus is on whether a potential digitally reachable donor would receive, notice, trust, respond to, and be practically able to act on an urgent request. By defining mobilisability when using digital tools, this study contributes to planning of donor-mobilisation interventions and systems that are technically feasible, behaviourally plausible, and locally appropriate.

## METHODS

### Study setting and design

We conducted a cross-sectional online survey among digitally reachable adults living in Ghana from 16 September 2025 to 31 January 2026. Eligible participants were aged 18 years or older and able to complete the survey online. The minimum required sample size was 384, calculated using Cochran’s formula with a 95% confidence level, a 5% margin of error and an assumed population proportion of 50% [31].

Recruitment was via targeted Facebook (Meta) advertisements across all 16 administrative regions, and non-probability sampling through collaborator and participant networks. Facebook was the most commonly reported recruitment source, accounting for 80% of participants. These approaches used established online recruitment methods used by members of the research team previously with Ukrainian refugees and also within Ghana [32,33]. Recruitment patterns were reviewed periodically, and later advertising campaigns focused on groups that were less represented in the accumulating sample, including women, old adults, residents of selected regions and people living in more rural areas.

The questionnaire (supplementary appendix 1) was administered through Qualtrics XM [34] in English, the official language of Ghana. The median recorded completion time was 18.9 min (IQR 14.4-25.7). It was co-developed with Ghanaian collaborators to ensure that the wording and content were appropriate to the local context. It contained up to 51 participant-facing questions covering sociodemographic characteristics (e.g. age, sex, location), mobile phone ownership and use, blood-donation knowledge and experience, donation concerns and practical barriers, trust in sources who may request blood, willingness to donate for different recipients, and anticipated blood-donation engagement. Participant information was provided prior to the start of the survey. Only participants who provided written informed electronic consent and confirmed that they were aged 18 years or older could continue. After completing the questionnaire, participants could voluntarily enter a prize draw. Twenty-five winners each received GH₵100 (approximately US$9) as an incentive for their time commitment.

Qualtrics duplicate-detection tools were used to identify repeated submissions, and internet protocol (IP) address-based geolocation was used to check that submissions originated in Ghana [35]. In total, 1,470 submissions were received. Responses were excluded if the questionnaire was incomplete, consent or age eligibility was not met, the submission was identified as a duplicate, or its location could not be verified as Ghana. Following screening, 1,067 valid responses from eligible participants who provided informed consent were retained for analysis.

### Outcome definition

Blood-donation research indicates that intention is an important antecedent of donation behaviour but does not alone capture whether donation will occur, with behavioural control and situational factors also influencing action [36]. Özcan et al.’s study of barriers to response in mobile-device-supported targeted responder systems, shows that digitally coordinated response may be constrained at multiple stages, including commitment, notification, leaving one’s current situation and performing the requested action [37]. Research on mobile blood-donation technologies has also examined willingness to install and use a blood-donation app separately from general donation intentions [22]. To capture anticipated engagement with digitally coordinated urgent blood-donor requests, we drew on these complementary lines of evidence to inform the conceptual framework shown in Figure 1 and defined digital urgent donor mobilisability as the primary analytical outcome. This was constructed as a binary formative indicator aligned with the study aim and intended to capture the perceived ability and willingness to act on urgent blood-donation requests sent through digital channels. Participants were coded as having high mobilisability if they met all four of the following criteria: 1) they were categorised as willing to donate blood in future, comprising those who answered “yes, very likely” or “possibly, if needed” (high future-donation willingness); 2) were likely or very likely to install a trusted blood-donation app (high trusted-app installation willingness); 3) were likely or very likely to respond and agree to donate after receiving a request from a trusted-source (high trusted-request response willingness); 4) and described themselves as somewhat or very flexible to leave their current activities to donate (high practical flexibility). Participants not meeting all four criteria were coded as not having high mobilisability.

**Figure 1:**
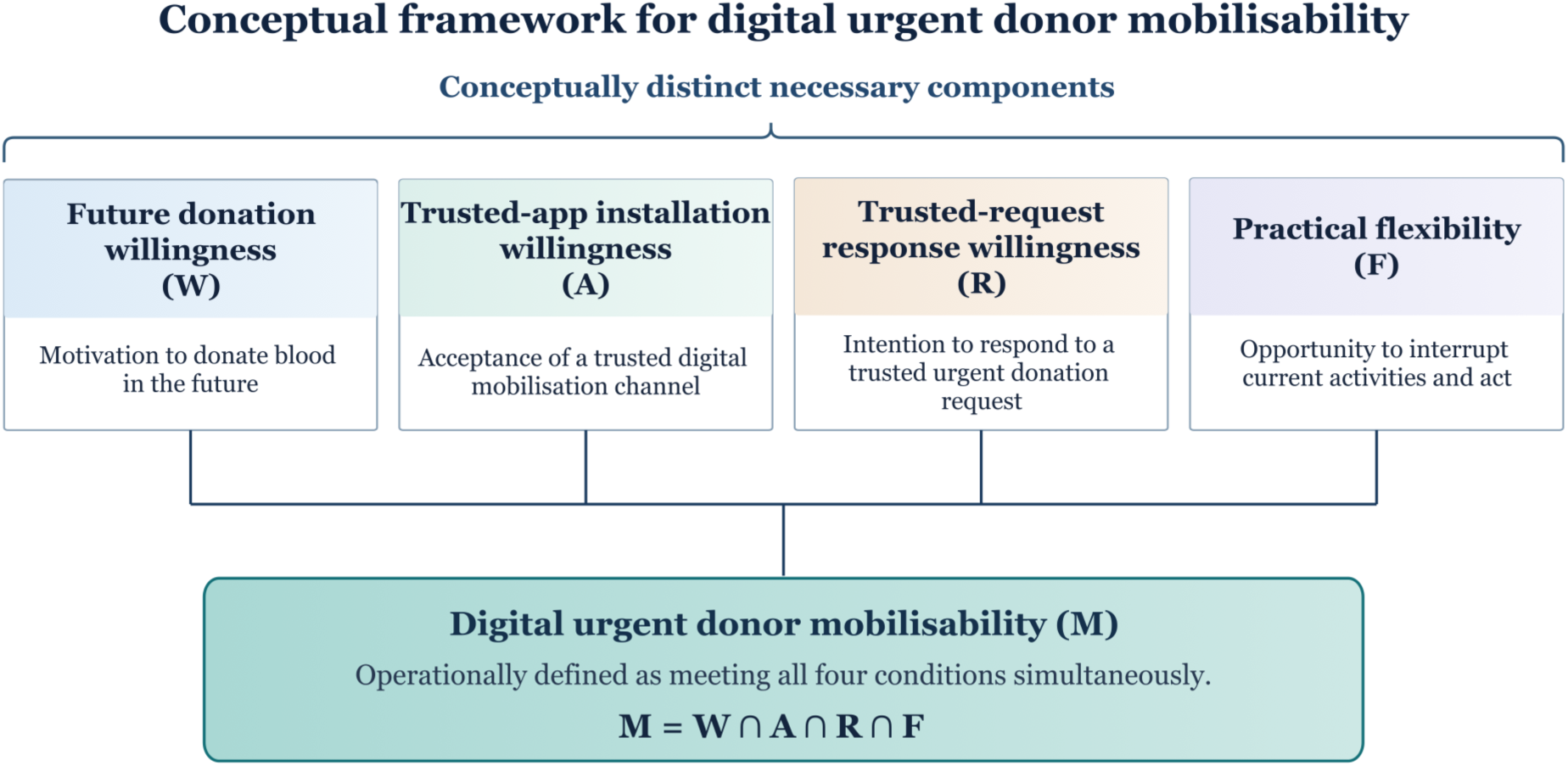
Conceptual framework for digital urgent donor mobilisability and its four-component operational definition. High mobilisability was defined as simultaneously meeting the criteria for high future donation willingness, high trusted-app installation willingness, high trusted-request response willingness, and high practical flexibility.

### Explanatory variables (predictors)

Sociodemographic variables included gender, age group, religion, ethnicity, region cluster, residence type, daytime/night-time travel time to nearest healthcare facility, education and economic activity status. Variables related to blood donation experience included previous donor status, blood-donation knowledge, exposure to blood-donation information, previous personal or close-network transfusion experience and donation incentive preference. Donation concerns were summarised as no concern, knowledge or eligibility concerns, fear or physical discomfort, religious or cultural concerns, health or safety concerns, and other concerns. Participants could select more than one concern domain. Donation concerns were not included in the primary model because no-concern status was structurally dependent on previous donor status: all participants classified as having no concerns were previous donors. Including both variables in the same model would therefore not provide independent adjustment. The effect of including this variable was examined in a sensitivity analysis. Practical barriers were represented by five binary indicators, each reflecting whether participants identified the following as a barrier to responding to an urgent blood-donation request: work schedule, school schedule, transport, family or childcare responsibilities, and weather.

High digital-response readiness captured whether participants had the practical digital conditions needed to receive and respond promptly to a request. Participants were classified as having high readiness if they met all three conditions: 1) they could install and use a new app without help or with minimal help; 2) were likely or very likely to answer a call identified as coming from a trusted source; 3) and usually checked an important notification within 30 seconds; and. Participants who did not meet all three conditions were classified as having low digital-response readiness.

Trust in blood-request sources was summarised into four binary domains. Formal health-system trust indicated high trust in a request from a hospital or clinic. Interpersonal network trust indicated high trust in at least one of two sources: family or friends, or other individual members of their community. Community or organisational trust indicated high trust in at least one of three sources: blood-donation groups (such as the National Blood Service), religious groups, or community leaders or local officials. Unknown individual trust indicated high trust in a request from a source not personally known to the participant. Responses of ‘somewhat’ or ‘strongly’ trust met the high-trust threshold.

Recipient-related donation willingness was summarised into three binary domains: 1) Socially-connected recipient willingness indicated high willingness to donate for at least one of the following: family or friends, personally admired public figures, or community members; 2) Anonymous stranger willingness indicated high willingness to donate for an unknown recipient when no additional details were provided; 3) Emotionally-prompted stranger willingness indicated high willingness to donate for an unknown recipient when the request included a message containing an urgent emotional appeal (eg, “a pregnant lady bleeding, can you give blood urgently?”). Responses of somewhat willing or strongly willing met the high-willingness threshold.

A detailed specification of each variable is described in supplementary appendix 2. All constructed domain-level variables were used in the primary model to avoid overparameterisation while preserving the conceptual content of the survey. Item-level variables were examined in sensitivity analyses (supplementary appendix 9-10).

### Statistical analysis

Descriptive statistics were used to summarise participant characteristics and mobilisability. Selected sample characteristics were descriptively compared with corresponding national distributions from Ghana’s 2021 Population and Housing Census [38,39]. Bivariable logistic regression was used to estimate crude ORs and 95% CIs for associations with high mobilisability. Variables with a variable-level likelihood-ratio test p<0.20 were considered for the primary multivariable model [40,41]. Residence, education and economic activity were retained a priori as sociodemographic covariates. The adjusted model reported AORs with 95% CIs. “Prefer not to say” and “I don’t know” responses were treated as missing. As complete data were available for 1,002/1,067 participants (93.9%) and no outcome data were missing, regression analyses were conducted using complete cases without imputation, given the limited loss of observations. Model discrimination was assessed using the area under the receiver operating characteristic curve (AUC), calibration using the Hosmer–Lemeshow test, and multicollinearity using variance inflation factors (VIF). All tests were two-sided, with p<0.05 considered statistically significant [42–44].

As secondary analyses, four multivariable logistic regression models treated the original mobilisability components as separate outcomes, using the same predictor set as the primary model. Two sensitivity analyses were conducted. The first examined specific donation concern types among participants reporting at least one concern and excluded previous donor status. The second replaced grouped digital-response readiness, trust-sources and recipient-willingness variables with their individual raw questionnaire items. Secondary and sensitivity analyses were exploratory. Analyses were performed using Python 3.12 with statsmodels 0.14.6. Reporting followed the STROBE guidelines for cross-sectional studies (supplementary appendix 12).

## RESULTS

### Participant characteristics and distribution of mobilisation outcomes

Table 1 summarises key characteristics of the 1,067 participants, and the prevalence of high mobilisability and its four components within participant subgroups. Regional distribution is summarised using the four region clusters shown in the table, with the full distribution across all 16 administrative regions provided in supplementary appendix 4. Most participants were male (67.0%), aged 35 years or younger (73.1%), educated to tertiary level (79.3%) and living in non-urban settings (60.1%). Previous blood donation was reported by 58.0%, and 64.1% had high digital-response readiness. Travel to the nearest healthcare facility took more than 30 min for 38.9% during the daytime and 42.3% at night. Overall, 577 participants (54.1%) met all four criteria for high digital urgent blood-donor mobilisability.

**Table 1.**
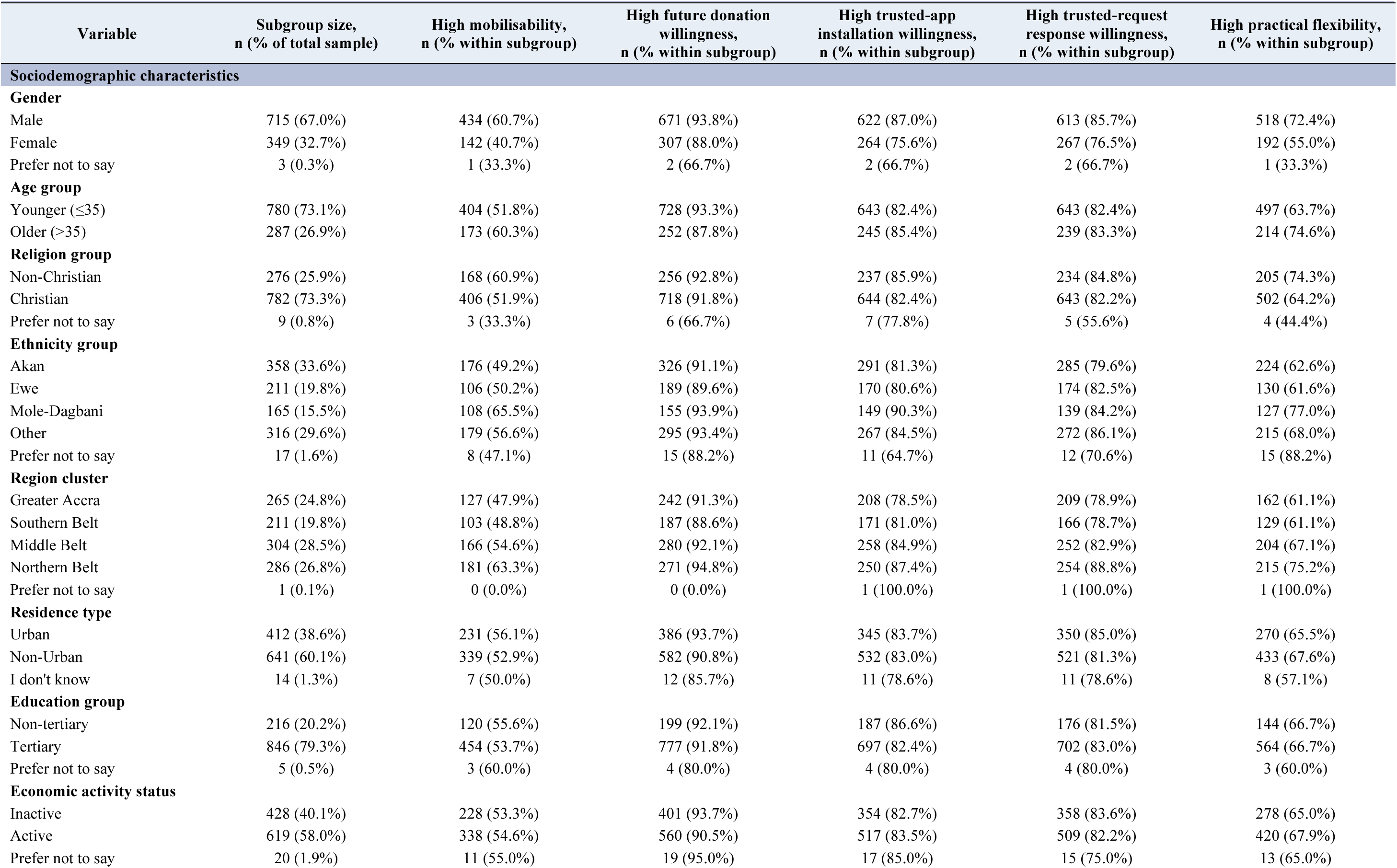
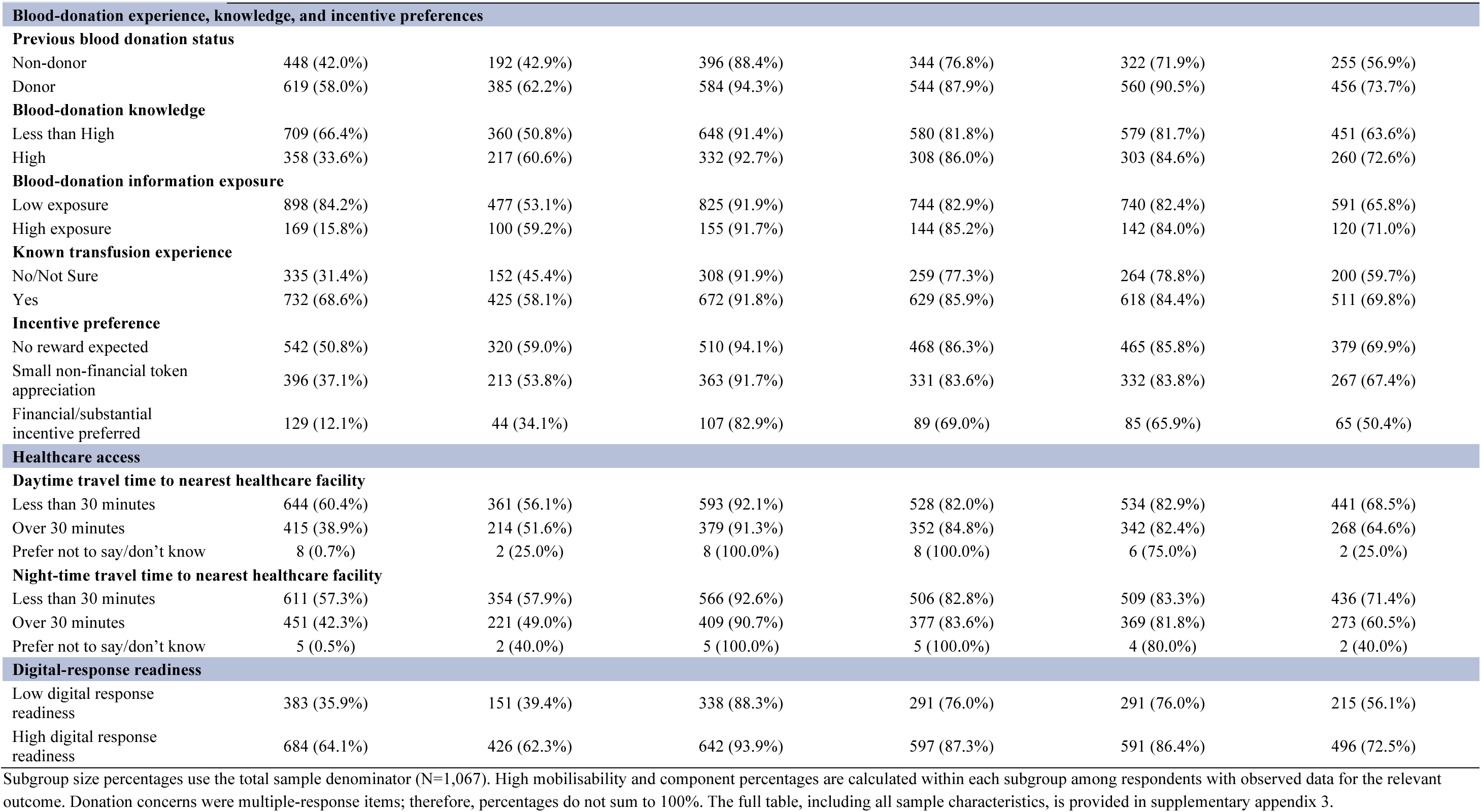
Selected sample characteristics and prevalence of high mobilisability and its four components (N=1,067)

Figure 2 shows the prevalence of participants meeting the predefined high threshold for each mobilisation component and the cumulative retention across the pathway. High future-donation willingness was reported by 91.8% of participants, while 83.2% reported high willingness to install a trusted app and 82.7% reported high willingness to respond to a trusted blood request. The proportion reporting high practical flexibility was comparatively lower at 66.6%. When the criteria were applied cumulatively, 79.2% met both the high future-donation willingness and high trusted-app installation willingness criteria. This fell to 71.7% after high trusted-request response willingness was added and to 54.1% after high practical flexibility was also required. Retention between successive stages was 86.2%, 90.5% and 75.4%, respectively. The largest reduction therefore occurred when high practical flexibility was added.

**Figure 2:**
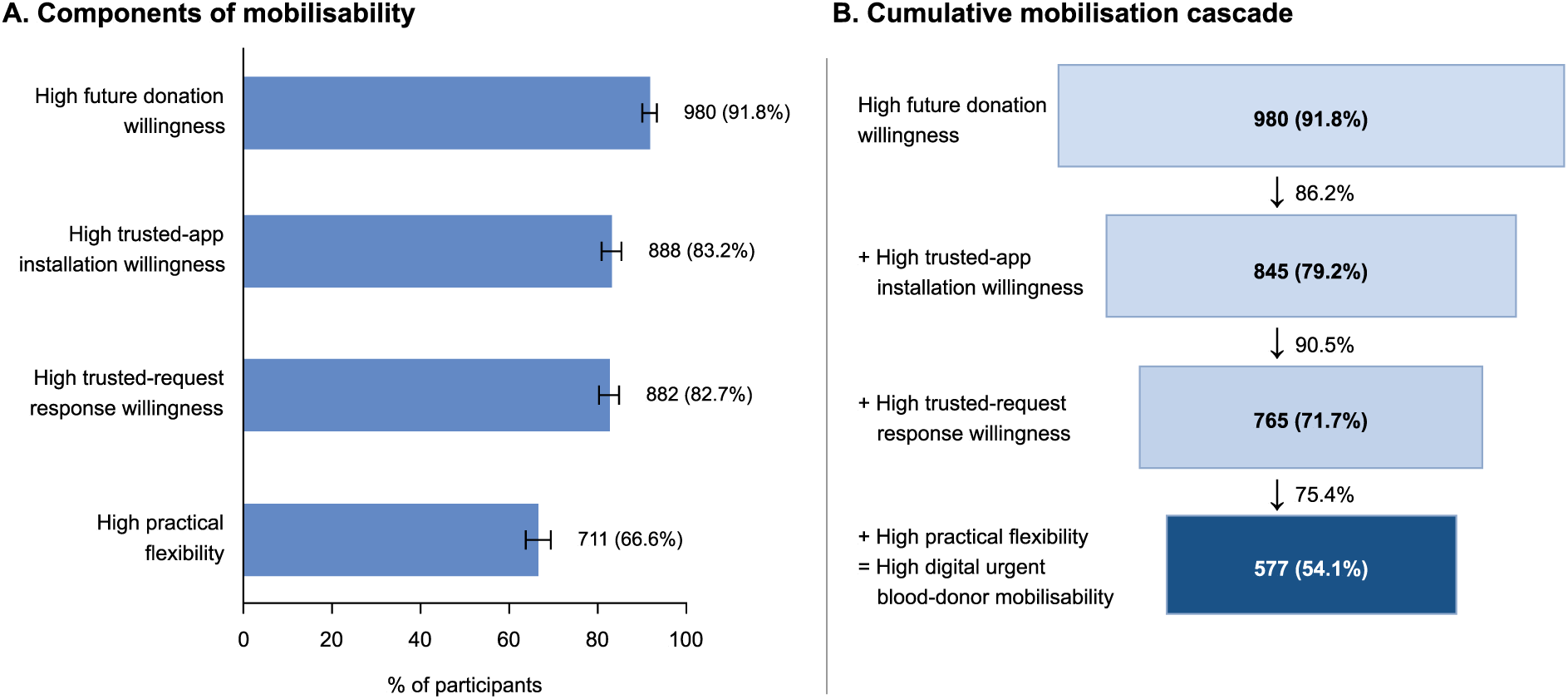
Mobilisability components and cumulative mobilisation cascade among survey participants (N=1,067). Panel A shows the proportion of participants meeting the predefined high threshold for each mobilisability component, with 95% confidence intervals (CIs). Panel B shows the cumulative proportion of the total analytical sample meeting the criteria at each successive stage, with percentages beside the downward arrows indicating retention from the preceding stage. The final stage represents high digital urgent blood-donor mobilisability.

### Factors associated with high digital urgent blood-donor mobilisability

Table 2 presents the crude and adjusted associations with high mobilisability. The primary model included 1,002 complete cases, of whom 548 (54.7%) had high mobilisability. Women had lower adjusted odds of high mobilisability than men (AOR 0.60, 95% CI 0.43–0.83). Mole-Dagbani participants had higher adjusted odds than Akan participants (AOR 1.96, 95% CI 1.08–3.58), and previous donors had higher adjusted odds than non-donors (AOR 1.47, 95% CI 1.08–2.01). Participants with high blood-donation knowledge also had higher adjusted odds than those with low knowledge (AOR 1.42, 95% CI 1.03–1.97).

**Table 2.**
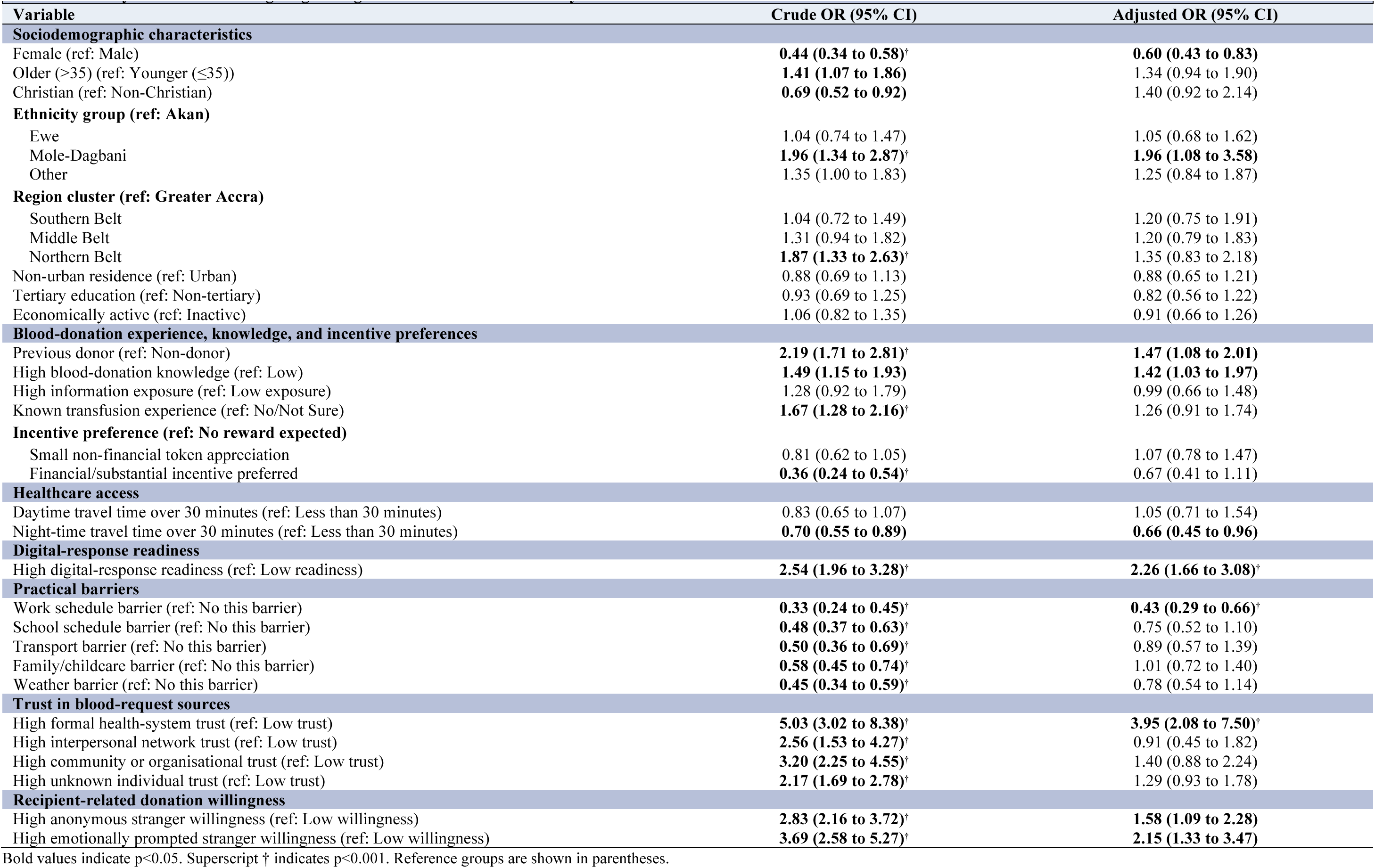
Primary determinants of high digital urgent blood-donor mobilisability.

|  |  |  |
| --- | --- | --- |
| **Variable** | **Crude OR (95% CI)** | **Adjusted OR (95% CI)** |
| **Sociodemographic characteristics** |  |  |
| Female (ref: Male) | **0.44 (0.34 to 0.58)<sup>†</sup>** | **0.60 (0.43 to 0.83)** |
| Older (>35) (ref: Younger (≤35)) | **1.41 (1.07 to 1.86)** | 1.34 (0.94 to 1.90) |
| Christian (ref: Non-Christian) | **0.69 (0.52 to 0.92)** | 1.40 (0.92 to 2.14) |
| **Ethnicity group (ref: Akan)** |  |  |
| Ewe | 1.04 (0.74 to 1.47) | 1.05 (0.68 to 1.62) |
| Mole-Dagbani | **1.96 (1.34 to 2.87)<sup>†</sup>** | **1.96 (1.08 to 3.58)** |
| Other | 1.35 (1.00 to 1.83) | 1.25 (0.84 to 1.87) |
| **Region cluster (ref: Greater Accra)** |  |  |
| Southern Belt | 1.04 (0.72 to 1.49) | 1.20 (0.75 to 1.91) |
| Middle Belt | 1.31 (0.94 to 1.82) | 1.20 (0.79 to 1.83) |
| Northern Belt | **1.87 (1.33 to 2.63)<sup>†</sup>** | 1.35 (0.83 to 2.18) |
| Non-urban residence (ref: Urban) | 0.88 (0.69 to 1.13) | 0.88 (0.65 to 1.21) |
| Tertiary education (ref: Non-tertiary) | 0.93 (0.69 to 1.25) | 0.82 (0.56 to 1.22) |
| Economically active (ref: Inactive) | 1.06 (0.82 to 1.35) | 0.91 (0.66 to 1.26) |
| **Blood-donation experience, knowledge, and incentive preferences** |  |  |
| Previous donor (ref: Non-donor) | **2.19 (1.71 to 2.81)<sup>†</sup>** | **1.47 (1.08 to 2.01)** |
| High blood-donation knowledge (ref: Low) | **1.49 (1.15 to 1.93)** | **1.42 (1.03 to 1.97)** |
| High information exposure (ref: Low exposure) | 1.28 (0.92 to 1.79) | 0.99 (0.66 to 1.48) |
| Known transfusion experience (ref: No/Not Sure) | **1.67 (1.28 to 2.16)<sup>†</sup>** | 1.26 (0.91 to 1.74) |
| **Incentive preference (ref: No reward expected)** |  |  |
| Small non-financial token appreciation | 0.81 (0.62 to 1.05) | 1.07 (0.78 to 1.47) |
| Financial/substantial incentive preferred | **0.36 (0.24 to 0.54)<sup>†</sup>** | 0.67 (0.41 to 1.11) |
| **Healthcare access** |  |  |
| Daytime travel time over 30 minutes (ref: Less than 30 minutes) | 0.83 (0.65 to 1.07) | 1.05 (0.71 to 1.54) |
| Night-time travel time over 30 minutes (ref: Less than 30 minutes) | **0.70 (0.55 to 0.89)** | **0.66 (0.45 to 0.96)** |
| **Digital-response readiness** |  |  |
| High digital-response readiness (ref: Low readiness) | **2.54 (1.96 to 3.28)<sup>†</sup>** | **2.26 (1.66 to 3.08)<sup>†</sup>** |
| **Practical barriers** |  |  |
| Work schedule barrier (ref: No this barrier) | **0.33 (0.24 to 0.45)<sup>†</sup>** | **0.43 (0.29 to 0.66)<sup>†</sup>** |
| School schedule barrier (ref: No this barrier) | **0.48 (0.37 to 0.63)<sup>†</sup>** | 0.75 (0.52 to 1.10) |
| Transport barrier (ref: No this barrier) | **0.50 (0.36 to 0.69)<sup>†</sup>** | 0.89 (0.57 to 1.39) |
| Family/childcare barrier (ref: No this barrier) | **0.58 (0.45 to 0.74)<sup>†</sup>** | 1.01 (0.72 to 1.40) |
| Weather barrier (ref: No this barrier) | **0.45 (0.34 to 0.59)<sup>†</sup>** | 0.78 (0.54 to 1.14) |
| **Trust in blood-request sources** |  |  |
| High formal health-system trust (ref: Low trust) | **5.03 (3.02 to 8.38)<sup>†</sup>** | **3.95 (2.08 to 7.50)<sup>†</sup>** |
| High interpersonal network trust (ref: Low trust) | **2.56 (1.53 to 4.27)<sup>†</sup>** | 0.91 (0.45 to 1.82) |
| High community or organisational trust (ref: Low trust) | **3.20 (2.25 to 4.55)<sup>†</sup>** | 1.40 (0.88 to 2.24) |
| High unknown individual trust (ref: Low trust) | **2.17 (1.69 to 2.78)<sup>†</sup>** | 1.29 (0.93 to 1.78) |
| **Recipient-related donation willingness** |  |  |
| High anonymous stranger willingness (ref: Low willingness) | **2.83 (2.16 to 3.72)<sup>†</sup>** | **1.58 (1.09 to 2.28)** |
| High emotionally prompted stranger willingness (ref: Low willingness) | **3.69 (2.58 to 5.27)<sup>†</sup>** | **2.15 (1.33 to 3.47)** |

Bold values indicate $p < 0.05$ . Superscript † indicates $p < 0.001$ . Reference groups are shown in parentheses.

Participants reporting more than 30 min of night-time travel to the nearest healthcare facility had lower adjusted odds of high mobilisability (AOR 0.66, 95% CI 0.45–0.96). Daytime travel exceeding 30 min was not associated with high mobilisability. Participants who regarded their work schedule as a barrier also had lower adjusted odds (AOR 0.43, 95% CI 0.29–0.66).

High trust in the health system showed the strongest positive adjusted association with high mobilisability (AOR 3.95, 95% CI 2.08–7.50). High willingness to donate to a stranger without additional information showed a positive association (AOR 1.58, 95% CI 1.09–2.28), while high willingness to donate to a stranger with an emotional message showed a stronger association (AOR 2.15, 95% CI 1.33–3.47). High willingness to donate to a socially-connected recipient was not included in the adjusted model because the low-willingness group contained no events. All participants with high mobilisability also described a high willingness to donate to socially-connected recipient.

High digital-response readiness was associated with more than twice the adjusted odds of high mobilisability (AOR 2.26, 95% CI 1.66–3.08). In a sensitivity analysis replacing this grouped measure with its individual items, high likelihood of answering a trusted-source call (AOR 2.65, 95% CI 1.64–4.30) and checking important notifications within 30 seconds (AOR 1.83, 95% CI 1.29–2.58) were positively associated with high mobilisability (supplementary appendix 10).

There were no clear adjusted associations with age, religion, region, residence, education or economic activity. The primary model had acceptable discrimination, with an AUC of 0.79. The Hosmer–Lemeshow p value of 0.24 provided no evidence of poor calibration. The maximum VIF of 2.29 indicated no evident multicollinearity.

### Component-specific associations across the mobilisation pathway

Figure 3 summarises selected adjusted associations across the four component outcomes, with complete estimates reported in the supplementary appendix 7. High digital-response readiness was positively associated with all four components (AORs 1.78–1.82). Women had lower odds of high future-donation willingness (AOR 0.48, 95% CI 0.28–0.85), high trusted app-installation willingness (AOR 0.53, 95% CI 0.35–0.80) and high practical flexibility (AOR 0.65, 95% CI 0.47–0.91). Adults aged >35 years had lower odds of high future-donation willingness (AOR 0.48, 95% CI 0.27–0.86) but higher odds of high practical flexibility (AOR 1.64, 95% CI 1.13–2.38).

**Figure 3:**
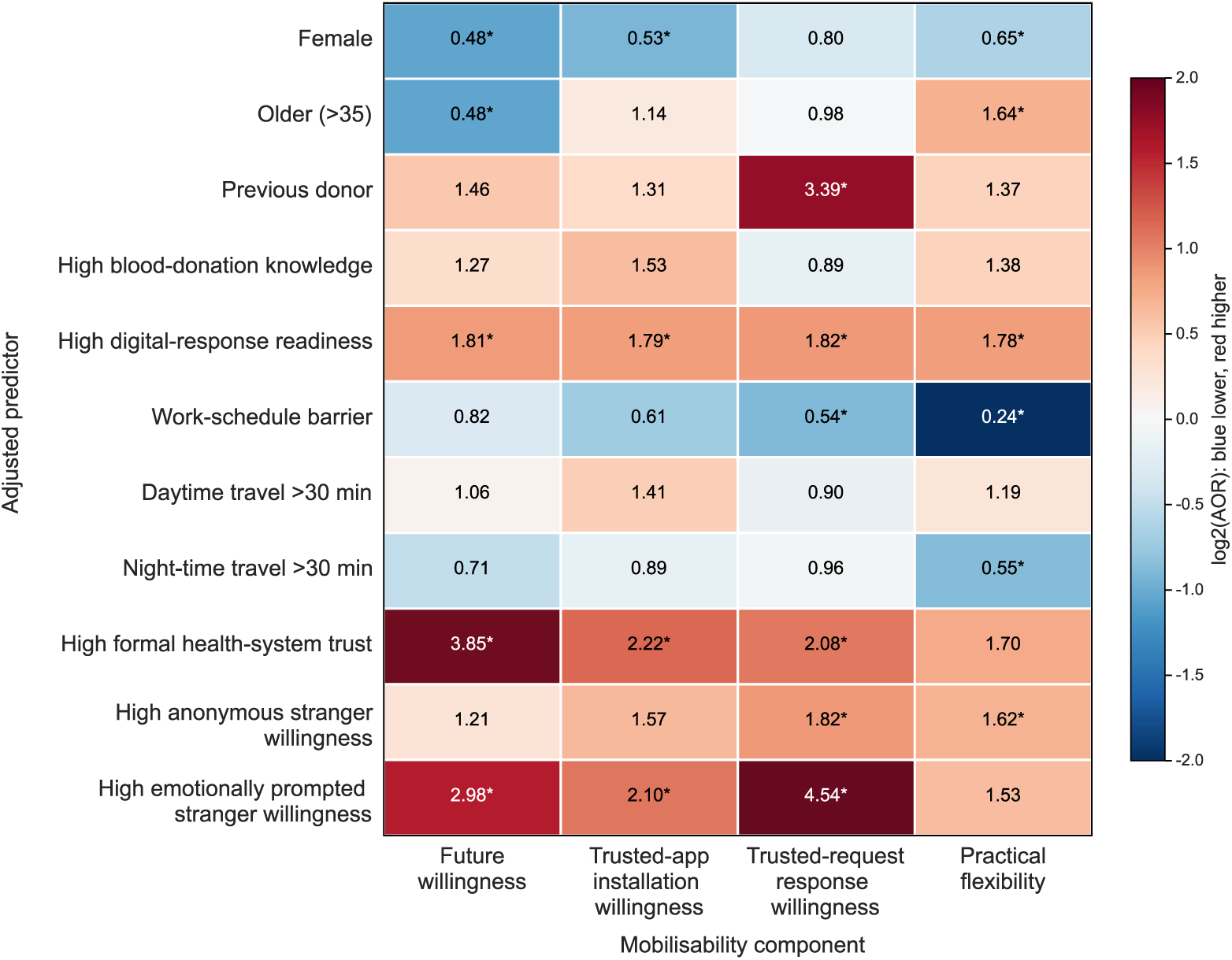
Adjusted associations between selected predictors and the four components of digital urgent blood-donor mobilisability. Cells show adjusted ORs (AORs) from separate component-specific multivariable logistic regression models. The colour scale represents the log2-transformed AOR, with values above 1 indicating higher odds and values below 1 indicating lower odds of meeting the component criterion. Asterisks indicate p<0.05. Complete model estimates are provided in supplementary appendix 7.

Previous donation experience was most strongly associated with high trusted-request response willingness (AOR 3.39, 95% CI 2.21–5.21). Night-time travel exceeding 30min was associated with lower odds of high practical flexibility (AOR 0.55, 95% CI 0.37–0.80). A work-schedule barrier was associated with lower odds of high trusted-request response willingness (AOR 0.54, 95% CI 0.29–1.00) and substantially lower odds of high practical flexibility (AOR 0.24, 95% CI 0.15–0.40). High formal health-system trust was not clearly associated with high practical flexibility but was strongly positively associated with the other three components. Emotionally prompted stranger willingness showed its strongest association with high trusted-request response willingness (AOR 4.54, 95% CI 2.67–7.71).

### Donation concerns and practical barriers across selected participant groups

Overall, 606 participants (56.8%) reported at least having one donation concern. Among these reporting any concern, health or safety concerns (47.0%), knowledge or eligibility concerns (46.2%) and fear or physical discomfort concerns (38.0%) were most common. Religious or cultural concerns were only reported by 33 participants (5.4%). Practical barriers were heavily reported by 992 participants (93.0%). Among these participants, transport was the most commonly reported barrier (88.2%), followed by work schedules (80.9%), weather (77.3%), school schedules (75.7%), and family or childcare responsibilities (59.6%).

Figure 4 shows the prevalence of donation concerns and practical barriers across selected participant groups. There were few concerns among those who had high mobilisability and a history of prior donation. High-mobilisability non-donors more frequently reported knowledge or eligibility concerns (57%), health or safety concerns (42%) and fear or physical discomfort concerns (35%).

**Figure 4:**
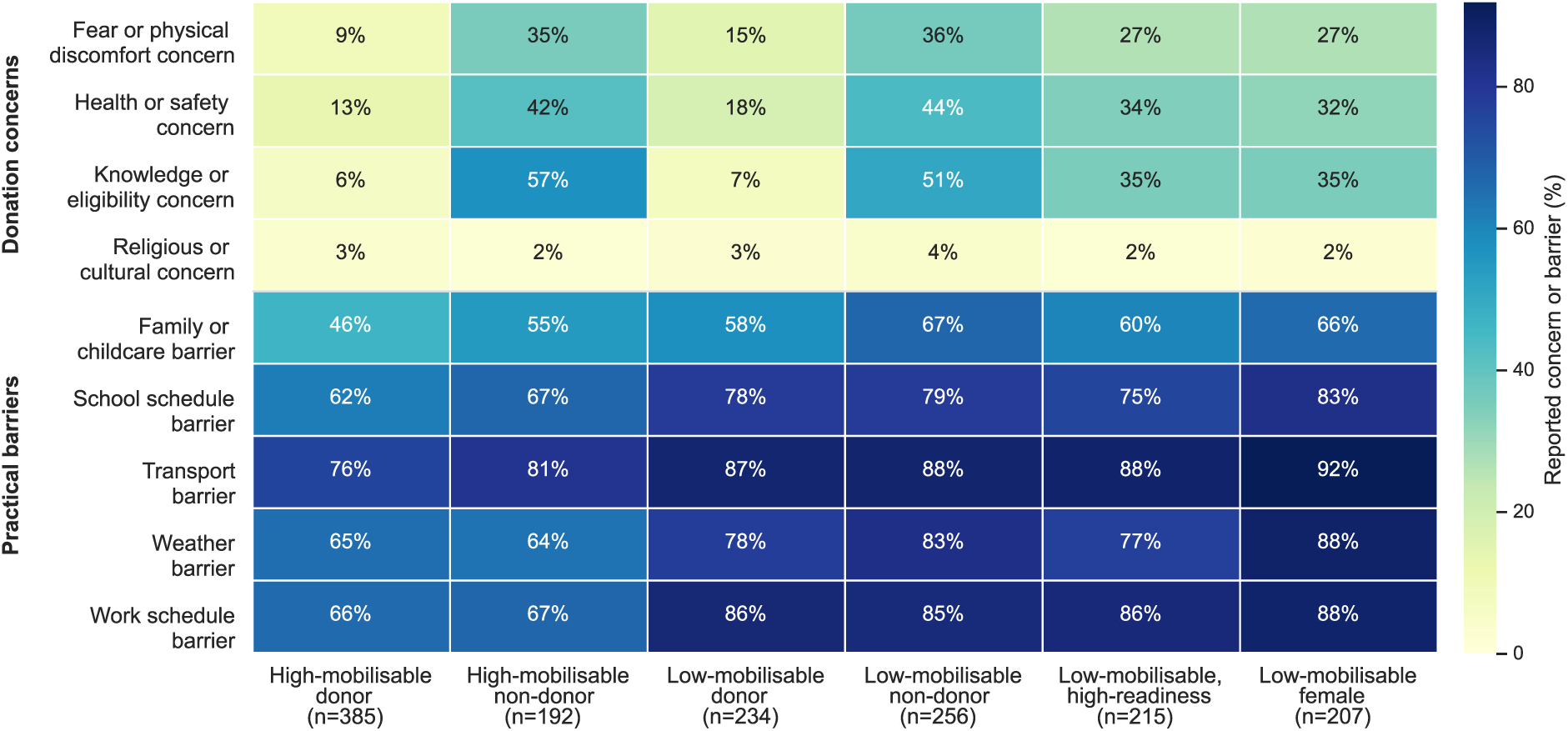
Prevalence of donation concerns and practical barriers across selected participant groups. Cells show the percentage of participants within each group reporting the corresponding concern or barrier. Participants could report more than one concern or practical barrier; therefore, percentages within groups do not sum to 100%. Group names and sizes are shown along the horizontal axis.

Practical barriers were more common in the low-mobilisability groups. Among low- mobilisability females, for example, 92% reported transport as a barrier, 88% reported weather and work schedules as barriers, 83% reported school schedules as a barrier, and 66% reported family or childcare responsibilities as a barrier. Among participants with high digital-response readiness, but low-mobilisability, work-schedules and transport were both reported as barriers by over 85%.

In the sensitivity analysis incorporating donation concern types into the model, most concern types did not show a statistically significant association with high mobilisability (supplementary appendix 9). Among practical barriers, work schedules remained associated with lower odds of high mobilisability (AOR 0.45, 95% CI 0.26–0.78), consistent with the primary model.

## DISCUSSION

Within the context of our study, digital reachability should not be equated with urgent blood donor mobilisability. Although willingness was overall high, only 54.1% of participants additionally reported sufficient practical flexibility to meet the definition of high mobilisability. Trust in the health system, and digital-response readiness, showed the strongest positive associations. Work-schedule barriers, longer night-time travel and female gender were associated with lower mobilisability. Component-specific models showed that these factors acted at different points in the digital urgent blood-donor mobilisation pathway.

The study extends previous blood-donation research in Ghana and sub-Saharan Africa, which has largely examined general willingness for routine donations [16–19]. Digital blood-donation studies have similarly focused on app acceptance, reminders, recruitment and routine engagement [22,26,45]. Our findings add an operational distinction important for urgent or emergency response and for resource planning in low resource setting: a person may be willing, digitally-connected and receptive to a trusted-request, yet still be unable to act at short notice given practical barriers. App adoption or message reach may therefore overestimate the pool of people available. Digital blood-donation programmes should be evaluated against the level of urgent need for blood, emotionally triggered messaging, and potential for timely arrival of a donor, rather than donors’ digital registrations or stated willingness alone.

Digital-response readiness was positively associated with all four components. Item-level analysis further suggested that answering trusted-source calls and checking important notifications promptly may be particularly important for translating digital reachability into the capacity to respond to urgent-requests. This extends previous evidence that usability, communication quality and integration with blood-service workflows influence the effectiveness of digital blood donation interventions [22,45], by identifying timely responsiveness as an additional requirement in urgent settings. Digital mobilisation systems could therefore assess potential donors’ response readiness and identify those with a high likelihood of noticing and acknowledging urgent requests promptly.

Formal health-system trust had the largest adjusted association with high mobilisability and was positively associated with future donation willingness, trusted-app installation willingness and trusted-request response willingness. This aligns with evidence that trust in blood-service institutions and the credibility of blood-donation message sources can influence donation intentions, while Ghanaian evidence has linked uncertainty about or lack of trust in what happens to donated blood with lower intention to return among first-time donors [46–48]. Requests should therefore come from identifiable hospitals or blood services, allow simple verification, and provide transparent information about how donated blood is managed and used to improve trustworthiness. Because trust in digital systems also depends on how information and requests are governed, transparent data-protection, request-authorisation and accountability procedures are also essential. This is also reinforced by previous research identifying privacy, misinformation and unequal access as concerns in digital health [49].

Work-schedule barriers were associated with lower mobilisability, lower trusted-request response willingness and particularly lower practical flexibility. Previous research in Ghana and sub-Saharan Africa has also identified time, inconvenience and distance as barriers to blood donation in routine settings [16,18,48]. Longer night-time, but not daytime, travel was also associated with lower mobilisability and practical flexibility. Data from Ghana’s 2021 Population and Housing Census document substantial inequalities in proximity to healthcare facilities, while other studies in Ghana show that longer travel times can reduce the use of time-sensitive health services [39,50]. Mobile blood-collection models have been used to improve access and collection capacity in other settings, and could also be complemented by innovations further along the blood-supply pathway. In Ghana, drone-based aerial logistics, such as Zipline, have been used to deliver blood products and other medical supplies rapidly to health facilities, with evidence from the Ashanti Region indicating improved timeliness and access to emergency commodities. However, their applicability and value for urgent blood-product distribution specifically, including optimal targeting and long-term sustainability, require further evaluation [51–53].

Women had lower mobilisability and lower donation willingness, trusted-app installation willingness and practical flexibility, but not lower trusted-request response willingness. This reinforces the male predominance reported among donors in Ghana and wider sub-Saharan Africa [16–19,48]. It suggests that women who receive a credible request may be willing to respond, but face different barriers to men elsewhere in the pathway, for example use of transport, work, weather, school and family or childcare barriers. Clinical eligibility is an important consideration. For example, prospective donors with haemoglobin below the required threshold are deferred from donation; women comprised most low-haemoglobin deferrals in a recent Ghanaian study [48,54]. Gender-responsive interventions could therefore consider inclusive approaches to urgent blood donation needs, for example with flexible collection, safe transport, support for caring responsibilities and appropriate eligibility and haemoglobin guidance.

Previous donors and people with high blood-donation knowledge had higher mobilisability. Familiarity with donation may reduce uncertainty and make a rapid decision easier, supporting the use of consent-based previous-donor registries for urgent activation [48]. However, 192 non-donors also met high mobilisability criteria. These participants frequently reported knowledge or eligibility, health or safety, and fear-related concerns. Concerns therefore did not always indicate a likely refusal to donate. A two-pathway model could rapidly contact eligible previous donors while providing mobilisable non-donors with eligibility guidance and first-donation support, thereby expanding and diversifying the future donor pool.

Willingness to donate for socially-connected recipients was almost universal in this sample, suggesting that existing relationships provide a strong motivation to help. This may also reflect local blood-donation practices, where patients in need of blood are often encouraged to seek potential donors among family and friends, particularly in urgent situations. Donation for strangers, by contrast, may depend more on need-based altruism (helping someone because they are in need, irrespective of personal ties) [55]. In our study, willingness to donate to strangers when the request included an emotional message appeared more strongly associated with high mobilisability, particularly trusted-request response, than willingness to donate to anonymous strangers without such framing. This may reflect empathy or compassion generated when the recipient’s need is made more concrete. Experimental studies have similarly found that recipient-focused emotional narratives and loss-framed emergency messages can increase donation intention or immediate willingness to donate [56,57]. Digital urgent requests could therefore include a brief and truthful account of why blood is needed. However, emotional content should remain proportionate and non-coercive, while any patient information should be used only with informed consent and appropriate privacy protection [58,59].

### Strengths and limitations

This study introduces an operational definition of digital urgent blood-donor mobilisability, combining willingness to donate, willingness to install a trusted-app, willingness to respond to a trusted urgent request and practical flexibility to act. This goes beyond measuring general donation intention or app acceptance by examining whether potential donors appear able to progress through the multiple stages required for urgent blood-donor mobilisation. The study also examined each component separately and considered demographic, digital, trust-related, donation-related, travel and practical factors. This allowed us to distinguish factors related to motivation, response to digital requests and the ability to act. The inclusion of participants from across Ghana, together with component-specific and sensitivity analyses, further strengthened the interpretation of the findings.

Compared with Ghana’s 2021 census, our participants were more likely to be male, aged 18–35 years and tertiary educated, while Christian affiliation and economic activity were closer to national distributions [38]. Detailed comparisons are provided in supplementary appendix 11. These patterns align with official evidence that women, older adults, rural residents and less-educated people experience greater digital exclusion in Ghana [60]. The sample may therefore represent a plausible early-user population for digital donor mobilisation, but the high prevalence of several outcome components should not be interpreted as nationally representative. The higher adjusted odds of high mobilisability among Mole-Dagbani compared to Akan participants should also be interpreted cautiously, as this may reflect sample composition or residual confounding and requires confirmation in further studies.

Other limitations should also be considered. As a cross-sectional observational study, the analysis cannot establish causality. Non-probability, English-language online recruitment also limits generalisability and may have under-represented people with limited internet access, lower literacy or less confidence using digital technologies. The measures were self-reported and captured anticipated rather than observed responses, which may be affected by social desirability and may not translate into actual attendance or completed donation during an emergency. Future studies could complement online recruitment with facility-based face-to-face data collection among potential donors and patients to explore whether findings differ across recruitment and survey modes. Finally, the mobilisability thresholds were defined for this study and require validation using prospective data from real-world urgent donor mobilisation systems.

## CONCLUSION

This study shows that digital reachability and donation willingness do not necessarily translate into urgent donor mobilisability. Effective mobilisation depends on trusted institutional requests, timely digital responsiveness and the practical ability to act. Work-schedule and night-time travel constraints may prevent otherwise willing individuals from responding promptly. Digital systems should therefore consider clearly identifiable and verifiable request sources, assess donors’ response readiness and current availability, and connect them with accessible collection options. Previous donors may provide an immediately responsive group, while highly mobilisable non-donors could expand the donor pool if given appropriate eligibility information and first-donation support. Prospective studies are needed to validate this mobilisability measure and determine whether these strategies improve timely attendance and completed donation during real-world blood shortages.

## DECLARATIONS

### Ethics approval

The study received ethics approval from the University of Southampton Ethics Committee, UK (ERGO number: 104098), and the University of Health and Allied Sciences Research Ethics Committee, Ghana (reference number: UHAS-REC A.9 [8] 24-25). All participants provided written informed consent online before they were able to access the questionnaire, and confirmed that they were aged 18 years or older. Participation was voluntary, and participants could exit the survey at any time before submission. The final analytic dataset was de-identified. No names, phone numbers or exact addresses were collected. Email addresses voluntarily provided for prize-draw administration or future follow-up research were stored separately and were not linked to survey responses.

### Patient and public involvementz

Patients and/or the public were not involved in the design, or conduct, or reporting, or dissemination plans of this research.

### Authorship contributions

HS, MH and MB co-led the conception and design of the study and developed the initial questionnaire. MAA and IAA contributed to the design and contextual adaptation of the questionnaire for Ghana, supported the ethics application and approval process in Ghana, and supported survey dissemination and recruitment. HS led the data collection, data cleaning and analysis, and preparation of the original manuscript draft. MH, MB, AC, MAA and IAA contributed to the interpretation of the findings and critically revised the manuscript for important intellectual content. All authors reviewed and approved the final manuscript for submission.

### Funding

This research is funded by the National Institute for Health and Care Research (NIHR) Biomedical Research Centre: Southampton. The views expressed are those of the author(s) and not necessarily those of the NIHR or the Department of Health and Social Care. HS is funded by the National Institute for Health and Care Research (NIHR) Biomedical Research Centre: Southampton.

## Acknowledgements

We acknowledge the support of Dr. Hintermann K.K. Mbroh, Chief Executive Officer, and his team at the Ho Teaching Hospital. We also acknowledge all those who gave their thoughts at both Ho and Hohoe hospitals. ChatGPT was solely used to assist with language editing and refinement of the manuscript’s wording. The entire manuscript including scientific content, analyses and interpretations was determined and reviewed by the human authors.

## Data availability statement

De-identified data underlying the findings of this study are available from the corresponding author upon reasonable request.

## Competing interests

None declared.

## Notes

### Competing Interest Statement

The authors have declared no competing interest.

### Summary of Updates

The manuscript title has been revised to "Beyond willingness to donate: a cross-sectional online survey to understand urgent blood-donor mobilisability among digitally reachable adults in Ghana" to better reflect the study's focus. The abstract has been shortened and restructured under the headings Objectives, Background, Methods, Results and Conclusion. The core content of the manuscript, including the study findings and conclusions, remains unchanged.

