## Supplementary appendix for "Beyond willingness to donate: a cross-sectional online survey to understand urgent blood-donor mobilisability among digitally reachable adults in Ghana"

### Supplementary material

#### Appendix 1: Survey instrument

**Participant Information and Consent (prior to beginning the online survey)**

This study has received ethics approval from the University of Southampton, UK (ERGO: 104098) and from the University of Health and Allied Sciences, Ghana (REC ref: UHAS-REC A.9 [8] 24-25).
Please read the following information before deciding to participate. You will need to confirm that you understand it to continue. At the end, you will be able to agree to take part in this survey.

**What is the research about?**

Hospitals frequently need blood donations urgently. Bleeding in pregnancy or road traffic incidents are common causes in Ghana for patients to need blood. Our study focuses on gathering and analysing Ghanaians' knowledge, attitudes, and experiences of blood donation, as well as their mobile phone usage patterns. By understanding these perspectives, the study aims to identify factors influencing individuals' willingness to donate and explore how digital health tools—such as mobile apps—could improve blood donation rates in urgent or emergency situations. This research is part of a project between the University of Southampton (UoS), UK and the University of Health and Allied Sciences (UHAS), Ghana. The ideas for this work came from UHAS, and with support from University of Southampton, we aim to generate practical insights that will benefit the health of people in Ghana.

**Why have I been asked to participate?**

You have been invited to take part because you are an adult living in Ghana and may have experiences or insights relevant to this research. We are seeking a range of participants from different regions and backgrounds. Your involvement is entirely voluntary and you may choose whether or not to participate.

**What will happen to me if I take part?**

If you decide to take part, you will be invited to complete a one-time online questionnaire, which should take around 15 minutes. You can fill it out on your own device (such as a smartphone or computer) at a time that suits you. There are no further requirements: no in-person visits, medical tests, or audio/video recordings. Once you have completed the questionnaire, you will have the option to provide your email address if you wish to be contacted for future, separate research projects—though that is entirely optional. Otherwise, there will be no follow-up after you finish. If you change your mind at any stage while completing the questionnaire, you can simply close it without penalty.

**Are there any benefits in my taking part?**

While there are no direct personal benefits for participating in this study, your input will help build a clearer understanding of the challenges surrounding blood donation in Ghana. In turn, this insight could guide the development of digital solutions that strengthen local healthcare systems and help save lives.

**Are there any risks involved?**

There are no anticipated risks involved in this study. The questionnaire will include questions about general demographic details (such as your region, education, occupation, and mobile phone use), as well as experiences and attitudes related to blood donation. Although demographic information is collected, your participation is completely anonymous, and individual responses cannot be traced back to you. Your privacy will be fully protected. If you feel uncomfortable answering any question, you can continue without answering, or withdraw from the survey entirely at any time without any consequences.

**What data will be collected?**

The questionnaire will collect information about your background, including your age group, gender, ethnicity, the part of Ghana where you live, education level, occupation, religion, and how long it typically takes you to reach your nearest hospital,etc. Additionally, we will gather details about whether you own a mobile phone, your device type, how frequently you use your phone, whether you find it easy to install new apps, and the types of activities you usually do on your phone, etc. We will also ask questions regarding your knowledge, attitudes, and experiences with blood donation—such as whether you've donated before, your motivations or concerns about donating, and your likelihood of responding to a blood donation request,etc. No personally identifiable information (such as your name or exact address, or your phone number) will be collected. All data collected from your questionnaire responses will remain completely anonymous and cannot be traced back to you, so you can feel confident in answering truthfully. We encourage you to provide information that is genuine and truly reflective of your actual situation, as this will help contribute to improving healthcare outcomes in Ghana. If you choose to provide your email address to be contacted for future studies, this information will be securely stored separately from your questionnaire responses and your answers will not be linked back to your email. All collected data will be securely handled and stored in a password-protected and encrypted environment, accessible only by authorised members of the research team, and your responses will remain completely anonymous.

**Will my participation be confidential?**

Your participation and the information we collect about you during the course of the research will be kept strictly confidential. Only members of the research team and responsible members of the University of Southampton may be given access to data about you for monitoring purposes and/or to carry out an audit of the study to ensure that the research is complying with applicable regulations. Individuals from regulatory authorities (people who check that we are carrying out the study correctly) may require access to your data. All of these people have a duty to keep your information, as a research participant, strictly confidential. We will ensure that all electronic data from your questionnaire responses are stored on password-protected devices or secure servers. Only members of the core research team will have access to the raw data, and any reports or publications will present the findings in a summarised or anonymised format. We will not share individual-level responses with any third parties. If you choose to provide an email address for future contact, it will be stored separately from your questionnaire data to ensure that it remains unlinked and untraceable to your personal responses.

**Do I have to take part?**

No, it is entirely up to you to decide whether or not to take part. If you decide you want to take part, you will need to sign a consent form to show you have agreed to take part. In this study, you'll first complete an online consent form by agreeing to the statements. You can exit the questionnaire at any time without penalty.

**What happens if I change my mind?**

You have the right to change your mind and withdraw at any time without giving a reason and without your participant rights being affected. You can exit the questionnaire at any time without penalty. Once submitted, your anonymised responses cannot be traced back to you or removed, so please only proceed if you feel fully comfortable. If you withdraw from the study, we will keep the information about you that we have already obtained for the purposes of achieving the objectives of the study only.

**What will happen to the results of the research?**

Your personal details will remain strictly confidential. Research findings made available in any reports or publications will not include information that can directly identify you without your specific consent. Any findings from this study will be reported in a manner that does not identify individual participants. We may publish the results in scientific journals, as short ‘policy briefs’, or lay summaries, but only in anonymised, summarised form. The data itself will be stored securely and may be retained for 10 years in compliance with institutional guidelines, after which it will be destroyed or archived in an anonymous format. Should you wish to see a summary of the results once the study is complete, please let us know, and we will be happy to share any published or publicly available outcomes.

**Where can I get more information?**

If you would like to discuss this research further, please contact Honghui Shen who would be happy to answer your questions.

**What happens if there is a problem?**

If you have a concern about any aspect of this study, you should speak to the researchers who will do their best to answer your questions. You can contact Honghui Shen or his main supervisor Dr. Markus Brede or the primary UHAS liaison, for any concerns. If you remain unhappy or have a complaint about any aspect of this study, please contact the University of Southampton Head Research Ethics and Governance (023 8059 5058,) or the University of Health and Allied Sciences Research Ethics Committee. Thank you for taking the time to read the information sheet and considering taking part in the research. For more information about the Data Protection Privacy Notice, please refer to https://git.soton.ac.uk/hs5n22/pis.

**<u>About the prize draw</u>**

After you finish the survey, you may choose to enter a prize draw. If you choose to enter, you must provide an email address so we can contact you if you win. If you provide an invalid, false, or misspelt email we cannot contact you and you will be deemed to have forfeited your prize.

To help prevent fraud and malicious multiple submissions by the same person, this Qualtrics survey uses built-in Fraud Detection and we will also analyse submission metadata (such as IP address, user agent headers, etc) to identify and review potential duplicate responses. Any personally identifiable information (PII) such as IP addresses will be used only to detect and prevent malicious duplicate entries and to filter out responses from outside Ghana; these data will not be included in the final research dataset. All data retained in the final research dataset will be anonymised so that it is not possible to trace responses back to individual participants.

Entering the prize draw does not guarantee that you will win. You may increase your chance of winning by inviting friends to complete the survey using your referral code; each friend who completes the survey increases your entries up to a maximum of five friends. Please note that every person may submit the survey only once — multiple submissions by the same person are not permitted.

For further details about the prize draw please refer to: https://git.soton.ac.uk/hs5n22/prize-draw (please rely on the most recent information published on that site).

**<u>Consent to take part in the survey</u>**

Thank you for reading the above information sheet and considering taking part in this research. To start answering questions of the survey, please click below to indicate that you have read and understood the above information, are **aged 18 or over** and agree to take part in this survey.

---

Q1 **By ticking the Agree box, you consent to participate in this survey and acknowledge that you are aged 18 or over and have read, understood and agreed to the above description of the research.**

○ Agree

○ Disagree

[Survey logic: If “Disagree” is selected, the participant is taken to the end of the questionnaire and the survey is ended.]

---

**Section 1: Demographics & Basic Information**

Q2 **Which region of Ghana do you currently live in?**

*Please provide as much as possible, as this will help improve healthcare in your community. Your response is completely anonymous.*

▼ Ahafo [Response format: single-select drop-down menu]

*Skip To: Q6 If Q2 = I don't know or Prefer not to say*

Q3 **Which district do you currently live in?**
*Please provide as much as possible, as this will help improve healthcare in your district. Your response is completely anonymous.*

▼ Asunafo North Municipal

Q4 [Display condition: shown only if Q3 = Hohoe]
**Which community in Hohoe do you live in?**
*If you live in more than one community, please select the one where you primarily reside. Please provide as much as possible, as this will help improve healthcare in your community. Your response is completely anonymous.*

▼ Alavanyo Wudidi

Q5 [Display condition: shown only if Q4 = Other(Please specify in the next question)]
**If your community is not listed above, please specify it here.**
Enter N/A if you don't know, skip this question if you prefer not to say. *Please provide as much as possible, as this will help improve healthcare in your community. Your response is completely anonymous.*

________________________________________

Q6 **How would you describe the area where your community is located?**

- ○ Urban / city – a densely populated area with extensive built-up infrastructure, major services, and commercial centres
- ○ Peri-urban / suburban – an area on the outskirts of a city or large town, with mixed residential and semi-rural features
- ○ Town / small urban centre – a moderately populated settlement that serves as a local market or administrative hub but is not a major city
- ○ Rural / village – a sparsely populated area with mainly agricultural or undeveloped land, limited services, and small settlements
- ○ I don't know

Q7 **During the daytime (e.g., when you are at work or school), how long does it typically take you to travel to the nearest hospital?** Please consider your usual mode of transportation, without accounting for traffic congestion or bad weather conditions.

- ○ Less than 30 minutes
- ○ 30–60 minutes
- ○ 1–2 hours
- ○ More than 2 hours
- ○ I don't know
- ○ Prefer not to say

Q8 **During the nighttime (e.g., when you are at home), how long does it typically take you to travel to the nearest hospital?** Please consider your usual mode of transportation, without accounting for traffic congestion or bad weather conditions.

- ○ Less than 30 minutes
- ○ 30–60 minutes
- ○ 1–2 hours
- ○ More than 2 hours
- ○ Prefer not to say

Q9 **What is your birth year range?**

▼ 2005–2007 (Aged 18–20)

Q10 **What is your gender?**
Your response is anonymous. Choosing a specific answer helps us improve public health in Ghana.

- ○ Male
- ○ Female
- ○ Prefer not to say

Q11 **What is your ethnicity?**
Your response is anonymous. Choosing a specific answer helps us improve public health in Ghana.

- ○ Akan
- ○ Ewe
- ○ Ga-Adangbe
- ○ Mole-Dagbani
- ○ Guan
- ○ Gurma
- ○ Grusi
- ○ Mande
- ○ Other (please specify in the box below) ________________________________________________
- ○ Prefer not to say

Q12 **Which of the following best describes your origin?**
Your response is anonymous. Choosing a specific answer helps us improve public health in Ghana.

- ○ I am native to Ghana and currently live in my birth region
- ○ I am native to Ghana but moved to my current home from a different region
- ○ I was born outside Ghana and migrated from another country
- ○ I am a foreign national currently working or living in Ghana
- ○ Prefer not to say

Q13 **Which religious belief or group do you subscribe to?** Your response is anonymous. Choosing a specific answer helps us improve public health in Ghana.

- ○ No religious affiliation
- ○ African Traditional Religion
- ○ Christianity (e.g., Catholic, Protestant, Pentecostal, Charismatic)
- ○ Islam
- ○ Other (please specify in the box below) ________________________________________________
- ○ Prefer not to say

Q14 **What is your highest level of education?**
Your response is anonymous. Choosing a specific answer helps us improve public health in Ghana.

- ○ No formal schooling
- ○ Primary school (completed or partially)
- ○ Junior high school
- ○ Senior high school
- ○ Vocational or technical training
- ○ Tertiary education (college/university)
- ○ Prefer not to say

Q15 **What is your current primary occupation?**
If you have multiple roles, please select the one that is your primary source of support (i.e., your main source of income or livelihood). Your response is anonymous. Choosing a specific answer helps us improve public health in Ghana.

- ○ Professional/Technical
- ○ Administrative/Managerial
- ○ Clerical
- ○ Sales/Trade
- ○ Service
- ○ Agric/Ani. Husbandry/Forest/Fishing/Hunting
- ○ Production & related work
- ○ Artisans
- ○ Homemaker
- ○ Student
- ○ Unemployed/None
- ○ Other (please specify in the box below) ________________________________________________
- ○ Prefer not to say

---

**Section 2: Mobile Phone Ownership and Usage**

Q16 **Do you own a mobile phone?**
Your response is anonymous. Choosing a specific answer helps us improve public health in Ghana.

- ○ Yes, a smartphone (internet-capable)
- ○ Yes, a basic phone (calls and SMS only)
- ○ No, but I have access to a smartphone shared with family/friends
- ○ No, but I have access to a basic phone shared with family/friends
- ○ No, and I do not have access to a phone

*Skip To: Q25 If Q16 = No, and I do not have access to a phone*

Q17 **How likely are you to answer a phone call immediately if it is not from a family member or friend, but the caller ID clearly shows it is from a trusted source?**
A trusted source could include a local hospital, a health worker, or a community support line.

- ○ Very likely (I would answer right away if I recognize the source)
- ○ Likely (I usually answer such calls, though not always immediately)
- ○ Sometimes (I may answer depending on the timing or situation)
- ○ Unlikely (I often ignore or miss calls unless I know the person)
- ○ Very unlikely (I rarely answer calls from unknown numbers, even if labeled)

Q18 **When you hear your phone ringing, how long does it usually take you to answer the call?**

- ◯ Immediately (Within 10 seconds)
- ◯ Quickly (Within 10–30 seconds)
- ◯ After a short while (Around 30–60 seconds)
- ◯ After a few minutes or longer (More than 1 minute)
- ◯ I usually miss the call but call back after seeing it
- ◯ I usually miss the call and do not call back

Q19 **Which of the following activities do you often do on your phone? (Select all that apply)**

- ☐ Basic Communication (making/receiving calls, sending/receiving SMS, using WhatsApp or other messaging apps)
- ☐ Social Networking (e.g., using Facebook, Instagram, Twitter, TikTok to interact with friends or post/share content)
- ☐ Browsing or Reading News (online articles, newspapers apps, social media feeds for current affairs)
- ☐ Searching for Information (e.g., health queries, directions, job opportunities, research, or general knowledge)
- ☐ Entertainment (playing mobile games, watching/streaming videos, listening to music)
- ☐ Online Shopping (buying or selling goods via e-commerce apps or websites)
- ☐ Other (please specify in the box below) ________________________________________

Q20 **How often do you usually check your phone in a day?**

- ◯ Constantly (multiple times every hour)
- ◯ About every hour
- ◯ Every 2–3 hours
- ◯ A few times a day
- ◯ Once a day or less

Q21 [Display condition: shown only if Q16 = *Yes, a smartphone (internet-capable)* or *No, but I have access to a smartphone shared with family/friends*]
**How likely are you to open or read a push notification on your phone immediately after it appears?** Push notification is a short message that pops up on a phone to remind or inform users about something important.

- ◯ Very likely (I usually check as soon as I see a notification)
- ◯ Likely (I check most of them, though not always right away)
- ◯ Sometimes (I may wait until it's convenient for me)
- ◯ Unlikely (I often ignore or dismiss most push notifications)
- ◯ Very unlikely (I rarely read push notifications)

Q22 [Display condition: shown only if Q16 = *Yes, a smartphone (internet-capable)* or *No, but I have access to a smartphone shared with family/friends*]
**On average, how long does it usually take you to check a push notification from an app you consider important?**
Please estimate the time, assuming you have allowed notifications from that app.

- ○ Immediately (Within 10 seconds)
- ○ Quickly (Within 10–30 seconds)
- ○ After a short while (Around 30–60 seconds)
- ○ After a few minutes or longer (More than 1 minute)
- ○ I usually miss the notification but check it later
- ○ I usually miss the notification and do not check it

Q23 [Display condition: shown only if Q16 = *Yes, a smartphone (internet-capable)* or *No, but I have access to a smartphone shared with family/friends*]
**On average, how much time do you spend using your smartphone each day?**
Please include all activities, such as calling, messaging, browsing, watching videos, playing games, and using apps.

- ○ Less than 1 hour
- ○ 1–2 hours
- ○ 2–4 hours
- ○ 4–6 hours
- ○ 6–8 hours
- ○ More than 8 hours

Q24 [Display condition: shown only if Q16 = *Yes, a smartphone (internet-capable)* or *No, but I have access to a smartphone shared with family/friends*]
**How confident do you feel about installing and using a new smartphone app on your own?**

- ◯ I can install and use new apps without any help
- ◯ I can install and use new apps with minimal help
- ◯ I might need occasional guidance, but I can manage
- ◯ I usually need someone else to help me
- ◯ I cannot do it without significant assistance

**Section 3: General Knowledge and Experience of Blood Donation**

---

Q25 **How would you rate your overall knowledge about blood donation (e.g., eligibility, process, benefits)?**

- ◯ I know little about blood donation, its process, or benefits.
- ◯ I am aware of the basic idea of blood donation but know only a few simple facts.
- ◯ I understand the general process, eligibility, and benefits, though not in detail.
- ◯ I am well-informed about the procedures, requirements, and benefits of blood donation.
- ◯ I have in-depth knowledge about blood donation, including detailed procedures and eligibility criteria.

Q26 **How many times have you donated blood?**

- ○ 0 times
- ○ 1 time
- ○ 2–5 times
- ○ More than 5 times

*Skip To: Q30 If Q26 = 0 times*

Q27 **Which of the following best describes the type of blood donation you have participated in?**

- ○ Voluntary Donation: You donated blood willingly, without receiving any payment, and **not for a specific person you know**.
- ○ Family Replacement Donation: You donated blood specifically to replace blood used by a family member, friend, or other identified patient.
- ○ Paid Donation: You donated blood in return for monetary payment.

Q28 **Which of the following best describes your blood donation behavior?**

- ○ Regular Donor: I donate blood on a regular schedule (e.g., every few months).
- ○ Casual Donor: I donate blood occasionally when I have the opportunity, even if there is no urgent need.
- ○ Need-Based Donor: I donate blood only when I see someone in urgent need or during emergencies.
- ○ Infrequent Donor: I have donated blood once or very rarely, without a consistent pattern.

Q29 **Is there anything that worries you or makes you unsure about donating blood (or donating more often)?**

- ○ Yes
- ○ No

Q30 **Which of the following worries or concerns do you have?** **(Select all that apply)**

- ☐ Not sure if I am eligible to donate
- ☐ Don't know how or where to donate blood
- ☐ Fear of needles or pain
- ☐ Fear of fainting or feeling unwell at the sight of blood
- ☐ Religious or cultural beliefs
- ☐ Not enough time or too inconvenient
- ☐ Concerns about infection or safety
- ☐ Worried that donating blood might be bad for my health
- ☐ Other (please specify in the box below) ________________________________________________

Q31 [Display condition: shown only if Q26 = *0 times*]
Based on information from the World Health Organization and Ghana's Ministry of Health, donating blood is a simple act that can save lives, ensuring hospitals have the essential supplies they need during emergencies. It is generally safe and does not cause harm to the donor. It also offers donors a free basic health check and fosters a sense of community support.
**Considering these benefits, would you consider donating blood in the future?**
You can read more at:

- WHO | Blood products: Blood donation - Questions and answers
- National Blood Service Ghana | Five (5) reasons why you should donate blood

○ Yes, very likely

○ Possibly, if needed

○ Not sure

○ Probably not

○ No, definitely not

Q32 [Display condition: shown only if Q26 ≠ *0 times*]
**Based on your past experience of donating blood, how willing are you to donate again in the future?**

○ Yes, very likely

○ Possibly, if needed

○ Not sure

○ Probably not

○ No, definitely not

Q33 **Have you, a close family member, or a friend you personally know ever received a blood transfusion?**

- ○ Yes, I have
- ○ Yes, a close family member has
- ○ Yes, a friend has
- ○ No
- ○ Not sure

*Skip To: Q36 If Q33 = No or Not sure*

Q34 **How did that experience affect your attitude toward blood donation?**

- ○ Significantly more willing
- ○ Somewhat more willing
- ○ No change
- ○ Somewhat less willing
- ○ Significantly less willing

Q35 **Why did this experience shape your current attitude toward blood donation?**
Please explain in 1-2 simple sentences why your attitude either remained unchanged or shifted (more positive or negative) after this experience.

________________________________________________________________

Q36 **How frequently do you come across blood donation-related information, either by actively seeking or passively receiving it?**

- ○ Never
- ○ Rarely
- ○ A few times a year
- ○ Monthly
- ○ Weekly
- ○ Daily

Q37 **Where do you primarily get information about blood donation?**

- ○ I have never noticed or come across any blood donation-related information
- ○ SMS or email notifications
- ○ Word of mouth (friends, family, or colleagues, etc.)
- ○ Printed materials (e.g., posters, flyers, brochures)
- ○ Official hospital or blood bank websites
- ○ Blood donation apps
- ○ Social media platforms
- ○ Television or radio advertisements

Q38 **For each of the following sources, please rate your level of trust in a blood donation request coming from that source**

| | Strongly trust | Somewhat trust | Neither trust nor distrust | Somewhat distrust | Strongly distrust |
|---|---|---|---|---|---|
| An official hospital or clinic | ○ | ○ | ○ | ○ | ○ |
| A family member or close friend | ○ | ○ | ○ | ○ | ○ |
| An unknown individual | ○ | ○ | ○ | ○ | ○ |
| A personal acquaintance or community member | ○ | ○ | ○ | ○ | ○ |
| A non-hospital/community blood donation group (e.g., social media public account, website, or app) | ○ | ○ | ○ | ○ | ○ |
| A religious organization | ○ | ○ | ○ | ○ | ○ |
| Community leader or local official | ○ | ○ | ○ | ○ | ○ |

Q39 **Please rate the following individuals or groups based on how willing you would be to donate blood if they needed it from you.**

| | Strongly willing | Somewhat willing | Neither willing nor unwilling | Somewhat unwilling | Strongly unwilling |
|---|---|---|---|---|---|
| Family or close friends (Someone you know personally who needs blood) | ○ | ○ | ○ | ○ | ○ |
| Public figures or celebrities you know (individuals with whom you have a direct/indirect connection or regard) | ○ | ○ | ○ | ○ | ○ |
| Community members or neighbours (Local residents or people within your community) | ○ | ○ | ○ | ○ | ○ |
| Complete strangers need blood with no additional details provided | ○ | ○ | ○ | ○ | ○ |
| Complete strangers with a compelling, urgent story or emotional appeal (e.g., pregnant lady bleeding, can you give blood urgently?) | ○ | ○ | ○ | ○ | ○ |

Q40 **If you received an urgent request to donate blood — through any digital channel (e.g., phone call, SMS, app notification) — from a trusted source, how likely are you to respond and agree to donate?**
A trusted source may include your local hospital, health authority, health worker, or verified health app, etc.

○ Very likely (I would respond and donate as soon as I can)

○ Likely (I would probably donate, but it depends on timing or circumstances)

○ Not sure (I would need to think about it or ask someone first)

○ Unlikely (I don’t usually respond to donation requests)

○ Very unlikely (I would probably ignore the request)

Q41 **How would you describe your ability to leave your current activity (e.g., study, work, household duties, farming, etc.) to donate blood if you receive an urgent blood request from a trusted source?**

- ○ Very flexible – I can leave on very short notice without major issues.
- ○ Somewhat flexible – I can usually leave, but I need minimal arrangement or notice.
- ○ Neutral – It depends on the situation; some arrangement is required.
- ○ Somewhat inflexible – It's difficult to leave, though I might manage under pressing circumstances.
- ○ Very inflexible – It's nearly impossible for me to leave my duties during the day.

Q42 **For each of the following factors, please indicate how much of a barrier it is to leaving your current activity to donate blood.**

| | No Barrier | Slight Barrier | Moderate Barrier | Strong Barrier | Very Strong Barrier |
|---|---|---|---|---|---|
| University or college schedule (e.g., lectures, exams) | ○ | ○ | ○ | ○ | ○ |
| Work schedule (inflexible hours) | ○ | ○ | ○ | ○ | ○ |
| Distance/transportation challenges | ○ | ○ | ○ | ○ | ○ |
| Family or childcare responsibilities | ○ | ○ | ○ | ○ | ○ |
| Weather conditions (e.g., heavy rain, extreme heat, flooding) | ○ | ○ | ○ | ○ | ○ |

Q43 **Any other barriers you can think of that might prevent you from leaving your current activity to donate blood?**

________________________________

Q44 **How likely are you to install a “Blood Donation” app on your phone if it were free and provided or used by a trusted source, such as your local hospital or health authority?**

- ○ Very likely
- ○ Likely
- ○ Not sure
- ○ Unlikely
- ○ Very unlikely

Q45 **What are the main reasons you would or would not like to install a “Blood Donation” app?**

________________________________________________________________

Q46 **If a blood donation app or website were designed with fun features—like earning points, unlocking badges, or competing with friends (similar to playing simple games)—how much would these features motivate you to donate blood?**

- ○ Extremely motivating
- ○ Very motivating
- ○ Moderately motivating
- ○ Slightly motivating
- ○ Not at all motivating

Q47 **If the points, badges, or similar fun features were linked to real rewards—such as discounts, gifts, or special recognition—how much would that further motivate you to donate blood?**

- ○ Extremely motivating
- ○ Very motivating
- ○ Moderately motivating
- ○ Slightly motivating
- ○ Not at all motivating

Q48 **Do you expect or desire any kind of reward or incentive when donating blood?**

- ○ No, I am willing to donate without any reward because I genuinely want to help others.
- ○ I appreciate small tokens of gratitude (e.g., refreshments, T-shirt, or certificate).
- ○ I would prefer financial compensation or a more substantial incentive.

Q49 **What kind of incentive or benefit (if any) would encourage you to donate blood more often?**
If you have more than one, separate them with a semicolon ( ; ). Leave blank if none.

________________________________________________________________

Q50 **Any other points you'd like to share about blood donation or Ghana's current public health situation?** Your complaints, visions, doubts—anything is welcome. Your response is anonymous and cannot trace back to you.

________________________________________________________________

Q51 **How did you hear about this survey? (Select all that apply)**

- ☐ Friends or family
- ☐ Colleague or workplace communication
- ☐ University or school communication
- ☐ Facebook
- ☐ WhatsApp
- ☐ LinkedIn
- ☐ Instagram
- ☐ X (formerly Twitter)
- ☐ TikTok
- ☐ Reddit
- ☐ Discord
- ☐ Email (personal or institutional)
- ☐ Health facility or staff
- ☐ Community or religious organisation
- ☐ Poster/Flyer or QR code
- ☐ Website or search engine (e.g., Google)
- ☐ Other (please specify) ________________________________________________

#### Appendix 2: Detailed specification of predictor variables

**Table A1. Construction and coding of predictor variables from questionnaire responses.**

| Predictor variable | Questionnaire item(s) | Coding from questionnaire responses | Final analytical representation |
|---|---|---|---|
| **Sociodemographic characteristics** | | | |
| **Gender** | Q10 | Male → Male; Female → Female. "Prefer not to say" → missing. | Binary variable; reference category = Male. |
| **Age group** | Q9 | Birth-year/age-range response recoded as Younger (≤35 years) or Older (>35 years). | Binary variable; reference category = Younger (≤35 years). |
| **Religion group** | Q13 | Christianity → Christian. No religious affiliation, African Traditional Religion, Islam and Other → Non-Christian. "Prefer not to say" → missing. | Binary variable; reference category = Non-Christian. |
| **Ethnicity group** | Q11 | Akan, Ewe and Mole-Dagbani retained as separate categories. Ga-Adangbe, Guan, Gurma, Grusi, Mande and self-described Other → Other. "Prefer not to say" → missing. | Categorical variable (4 levels); reference category = Akan. |
| **Region cluster** | Q2 | Greater Accra retained as a standalone category. The remaining administrative regions were collapsed into the prespecified Southern Belt, Middle Belt and Northern Belt categories used in the analysis. "Prefer not to say"/unknown → missing. | Categorical variable (4 levels); reference category = Greater Accra. See the verification note below for the exact region-to-belt mapping. |
| **Residence type** | Q6 | Urban / city → Urban. Peri-urban / suburban, Town / small urban centre and Rural / village → Non-Urban. "I don't know" → missing. | Binary variable; reference category = Urban. |
| **Education group** | Q14 | Tertiary education (college/university) → Tertiary. All other reported education levels → Non-tertiary. "Prefer not to say" → missing. | Binary variable; reference category = Non-tertiary. |
| **Economic activity status** | Q15 | Respondents in any listed working occupation were classified as Active: professional/technical, administrative/managerial, clerical, sales/trade, service, agriculture/animal husbandry/forestry/fishing/hunting, production-related work, artisan work and other working occupations. Homemakers, students, and respondents who were unemployed or reported no occupation were classified as Inactive. "Prefer not to say" → missing. | Binary variable; reference category = Inactive. |
| **Blood-donation experience, knowledge and incentive preferences** | | | |
| **Previous blood donation status** | Q26 | 0 times → Non-donor. 1 time, 2–5 times or >5 times → Donor. | Binary variable; reference category = Non-donor. |
| **Blood-donation knowledge** | Q25 | The two highest response levels were classified as High. The first three response levels were classified as Low. | Binary variable; reference category = Low. |
| **Blood-donation information exposure** | Q36 | Monthly, weekly or daily → High exposure. Never, rarely or a few times a year → Low exposure. | Binary variable; reference category = Low exposure. |
| **Known transfusion experience** | Q33 | Any "Yes" response for self, a close family member or a friend → Yes. "No" or "Not sure" → No/Not sure. | Binary variable; reference category = No/Not sure. |
| **Incentive preference** | Q48 | Responses were grouped into three categories: No reward expected; Small non-financial token appreciation; Financial/substantial incentive preferred. | Categorical variable (3 levels); reference category = No reward expected. |
| **Healthcare access** | | | |
| **Daytime travel time to nearest healthcare facility** | Q7 | Less than 30 minutes → <30 minutes. 30–60 minutes, 1–2 hours or >2 hours → 30 minutes or longer. "I don't know"/"Prefer not to say" → missing. | Binary variable; reference category = <30 minutes. |
| **Night-time travel time to nearest healthcare facility** | Q8 | Less than 30 minutes → <30 minutes. 30–60 minutes, 1–2 hours or >2 hours → 30 minutes or longer. "Prefer not to say" → missing. | Binary variable; reference category = <30 minutes. |

| Predictor variable | Questionnaire item(s) | Coding from questionnaire responses | Final analytical representation |
|---|---|---|---|
| **Digital response readiness** | | | |
| **High digital response readiness** | Q17, Q22, Q24 | High readiness required all three criteria: (1) Q24: could install or use a new app without help or with minimal help; (2) Q17: likely or very likely to answer a call identified as coming from a trusted source; and (3) Q22: usually checked an important app notification within 30 seconds (immediately [<10 s] or quickly [10–30 s]). Respondents who did not meet all three criteria were classified as having lower digital response readiness. | Constructed binary variable (all three criteria required); reference category = lower digital response readiness. |
| **Donation concerns** | | | |
| **No concern** | Q29–Q30 | No donation concern was reported. Because Q29–Q30 were shown according to previous donation status, this variable was structurally dependent on previous donor status in the analysed data. | Binary indicator; not included in the same primary model as previous donor status because the two variables were structurally dependent. |
| **Knowledge or eligibility concern** | Q30 | Reported at least one of the following concerns: "Not sure if I am eligible to donate"; "Don't know how or where to donate blood". | Binary multiple-response variable; reference category = respondents who reported none of these concerns. |
| **Fear or physical discomfort concern** | Q30 | Reported at least one of the following concerns: "Fear of needles or pain"; "Fear of fainting or feeling unwell at the sight of blood". | Binary multiple-response variable; reference category = respondents who reported none of these concerns. |
| **Religious or cultural concern** | Q30 | Reported "Religious or cultural beliefs" as a concern. | Binary indicator; reference category = respondents who did not report this concern. |
| **Health or safety concern** | Q30 | Reported at least one of the following concerns: "Concerns about infection or safety"; "Worried that donating blood might be bad for my health". | Binary multiple-response variable; reference category = respondents who reported none of these concerns. |
| **Other concern** | Q30 | Reported a concern using the free-text "Other" option. | Binary indicator; reference category = respondents who did not report another concern using the "Other" option. |
| **Practical barriers** | | | |
| **Work schedule barrier** | Q42 – Work schedule | No Barrier → no work schedule barrier. Slight, Moderate, Strong or Very Strong Barrier → work schedule barrier. | Binary variable; reference category = no work schedule barrier. |
| **School schedule barrier** | Q42 – University/college schedule | No Barrier → no school schedule barrier. Slight, Moderate, Strong or Very Strong Barrier → school schedule barrier. | Binary variable; reference category = no school schedule barrier. |
| **Transport barrier** | Q42 – Distance/transportation | No Barrier → no transport barrier. Slight, Moderate, Strong or Very Strong Barrier → transport barrier. | Binary variable; reference category = no transport barrier. |
| **Family/childcare barrier** | Q42 – Family/childcare responsibilities | No Barrier → no family/childcare barrier. Slight, Moderate, Strong or Very Strong Barrier → family/childcare barrier. | Binary variable; reference category = no family/childcare barrier. |
| **Weather barrier** | Q42 – Weather conditions | No Barrier → no weather barrier. Slight, Moderate, Strong or Very Strong Barrier → weather barrier. | Binary variable; reference category = no weather barrier. |
| **Trust in blood-request sources** | | | |
| **Formal health-system trust** | Q38 – Official hospital or clinic | Strongly trust or Somewhat trust → high trust; all other responses → low trust. | Binary trust variable (single item); reference category = low trust in the formal health-system source. |
| **Interpersonal network trust** | Q38 – Family member/close friend; personal acquaintance/community member | High trust was assigned when Strongly trust or Somewhat trust was reported for at least one of the two sources; otherwise, low trust was assigned. | Constructed binary trust variable (at least one source required); reference category = low trust in interpersonal network sources. |
| **Community or organisational trust** | Q38 – Non-hospital/community blood-donation group; religious organisation; community leader/local official | High trust was assigned when Strongly trust or Somewhat trust was reported for at least one of the three sources; otherwise, low trust was assigned. | Constructed binary trust variable (at least one source required); reference category = low trust in community or organisational sources. |
| **Unknown individual trust** | Q38 – Unknown individual | Strongly trust or Somewhat trust → high trust in unknown individuals; all other responses → low trust in unknown individuals. | Binary trust variable (single item); reference category = low trust in unknown individuals. |
| **Recipient-related donation willingness** | | | |
| **Socially-connected recipient willingness** | Q39 – Family/close friends; public figures/celebrities known to respondent; community members/neighbours | High willingness was assigned when Strongly willing or Somewhat willing was reported for at least one of the three recipient types; otherwise, low willingness was assigned. | Constructed binary willingness variable (at least one recipient type required); reference category = low willingness for socially-connected recipients. |
| **Anonymous stranger willingness** | Q39 – Complete stranger, no additional details | Strongly willing or Somewhat willing → willing to donate blood for an anonymous stranger; all other responses → low willingness. | Binary willingness variable (single item); reference category = low willingness for an anonymous stranger. |

| Predictor variable | Questionnaire item(s) | Coding from questionnaire responses | Final analytical representation |
|---|---|---|---|
| **Emotionally prompted stranger willingness** | Q39 – Complete stranger with compelling urgent story/emotional appeal | Strongly willing or Somewhat willing → willing to donate blood for a stranger described using a compelling urgent story or emotional appeal; all other responses → low willingness. | Binary willingness variable (single item); reference category = low willingess for an emotionally prompted stranger. |

### Appendix 3: Full sample characteristics with complete list of variables

**Table A2. Full sample characteristics and prevalence of high mobilisability and its four components (N=1,067)**

| Variable | Subgroup size, n (% of total sample) | High mobilisability, n (% within subgroup) | High future donation willingness, n (% within subgroup) | High trusted app installation willingness, n (% within subgroup) | High trusted-request response willingness, n (% within subgroup) | High practical flexibility, n (% within subgroup) |
|---|---|---|---|---|---|---|
| **Sociodemographic characteristics** | | | | | | |
| **Gender** | | | | | | |
| Male | 715 (67.0%) | 434 (60.7%) | 671 (93.8%) | 622 (87.0%) | 613 (85.7%) | 518 (72.4%) |
| Female | 349 (32.7%) | 142 (40.7%) | 307 (88.0%) | 264 (75.6%) | 267 (76.5%) | 192 (55.0%) |
| Prefer not to say | 3 (0.3%) | 1 (33.3%) | 2 (66.7%) | 2 (66.7%) | 2 (66.7%) | 1 (33.3%) |
| **Age group** | | | | | | |
| Younger (≤35) | 780 (73.1%) | 404 (51.8%) | 728 (93.3%) | 643 (82.4%) | 643 (82.4%) | 497 (63.7%) |
| Older (>35) | 287 (26.9%) | 173 (60.3%) | 252 (87.8%) | 245 (85.4%) | 239 (83.3%) | 214 (74.6%) |
| **Religion group** | | | | | | |
| Non-Christian | 276 (25.9%) | 168 (60.9%) | 256 (92.8%) | 237 (85.9%) | 234 (84.8%) | 205 (74.3%) |
| Christian | 782 (73.3%) | 406 (51.9%) | 718 (91.8%) | 644 (82.4%) | 643 (82.2%) | 502 (64.2%) |
| Prefer not to say | 9 (0.8%) | 3 (33.3%) | 6 (66.7%) | 7 (77.8%) | 5 (55.6%) | 4 (44.4%) |
| **Ethnicity group** | | | | | | |
| Akan | 358 (33.6%) | 176 (49.2%) | 326 (91.1%) | 291 (81.3%) | 285 (79.6%) | 224 (62.6%) |
| Ewe | 211 (19.8%) | 106 (50.2%) | 189 (89.6%) | 170 (80.6%) | 174 (82.5%) | 130 (61.6%) |
| Mole-Dagbani | 165 (15.5%) | 108 (65.5%) | 155 (93.9%) | 149 (90.3%) | 139 (84.2%) | 127 (77.0%) |
| Other | 316 (29.6%) | 179 (56.6%) | 295 (93.4%) | 267 (84.5%) | 272 (86.1%) | 215 (68.0%) |
| Prefer not to say | 17 (1.6%) | 8 (47.1%) | 15 (88.2%) | 11 (64.7%) | 12 (70.6%) | 15 (88.2%) |
| **Region cluster** | | | | | | |
| Greater Accra | 265 (24.8%) | 127 (47.9%) | 242 (91.3%) | 208 (78.5%) | 209 (78.9%) | 162 (61.1%) |
| Southern Belt | 211 (19.8%) | 103 (48.8%) | 187 (88.6%) | 171 (81.0%) | 166 (78.7%) | 129 (61.1%) |
| Middle Belt | 304 (28.5%) | 166 (54.6%) | 280 (92.1%) | 258 (84.9%) | 252 (82.9%) | 204 (67.1%) |
| Northern Belt | 286 (26.8%) | 181 (63.3%) | 271 (94.8%) | 250 (87.4%) | 254 (88.8%) | 215 (75.2%) |
| Prefer not to say | 1 (0.1%) | 0 (0.0%) | 0 (0.0%) | 1 (100.0%) | 1 (100.0%) | 1 (100.0%) |
| **Residence type** | | | | | | |
| Urban | 412 (38.6%) | 231 (56.1%) | 386 (93.7%) | 345 (83.7%) | 350 (85.0%) | 270 (65.5%) |
| Non-Urban | 641 (60.1%) | 339 (52.9%) | 582 (90.8%) | 532 (83.0%) | 521 (81.3%) | 433 (67.6%) |
| I don't know | 14 (1.3%) | 7 (50.0%) | 12 (85.7%) | 11 (78.6%) | 11 (78.6%) | 8 (57.1%) |
| **Education group** | | | | | | |
| Non-tertiary | 216 (20.2%) | 120 (55.6%) | 199 (92.1%) | 187 (86.6%) | 176 (81.5%) | 144 (66.7%) |
| Tertiary | 846 (79.3%) | 454 (53.7%) | 777 (91.8%) | 697 (82.4%) | 702 (83.0%) | 564 (66.7%) |
| Prefer not to say | 5 (0.5%) | 3 (60.0%) | 4 (80.0%) | 4 (80.0%) | 4 (80.0%) | 3 (60.0%) |
| **Economic activity status** | | | | | | |
| Inactive | 428 (40.1%) | 228 (53.3%) | 401 (93.7%) | 354 (82.7%) | 358 (83.6%) | 278 (65.0%) |
| Active | 619 (58.0%) | 338 (54.6%) | 560 (90.5%) | 517 (83.5%) | 509 (82.2%) | 420 (67.9%) |
| Prefer not to say | 20 (1.9%) | 11 (55.0%) | 19 (95.0%) | 17 (85.0%) | 15 (75.0%) | 13 (65.0%) |
| **Blood-donation experience, knowledge, and incentive preferences** | | | | | | |
| **Previous blood donation status** | | | | | | |
| Non-donor | 448 (42.0%) | 192 (42.9%) | 396 (88.4%) | 344 (76.8%) | 322 (71.9%) | 255 (56.9%) |
| Donor | 619 (58.0%) | 385 (62.2%) | 584 (94.3%) | 544 (87.9%) | 560 (90.5%) | 456 (73.7%) |

| Variable | Subgroup size, n (% of total sample) | High mobilisability, n (% within subgroup) | High future donation willingness, n (% within subgroup) | High trusted app installation willingness, n (% within subgroup) | High trusted-request response willingness, n (% within subgroup) | High practical flexibility, n (% within subgroup) |
|---|---|---|---|---|---|---|
| **Blood-donation knowledge** | | | | | | |
| Less than High | 709 (66.4%) | 360 (50.8%) | 648 (91.4%) | 580 (81.8%) | 579 (81.7%) | 451 (63.6%) |
| High | 358 (33.6%) | 217 (60.6%) | 332 (92.7%) | 308 (86.0%) | 303 (84.6%) | 260 (72.6%) |
| **Blood-donation information exposure** | | | | | | |
| Low exposure | 898 (84.2%) | 477 (53.1%) | 825 (91.9%) | 744 (82.9%) | 740 (82.4%) | 591 (65.8%) |
| High exposure | 169 (15.8%) | 100 (59.2%) | 155 (91.7%) | 144 (85.2%) | 142 (84.0%) | 120 (71.0%) |
| **Known transfusion experience** | | | | | | |
| No/Not Sure | 335 (31.4%) | 152 (45.4%) | 308 (91.9%) | 259 (77.3%) | 264 (78.8%) | 200 (59.7%) |
| Yes | 732 (68.6%) | 425 (58.1%) | 672 (91.8%) | 629 (85.9%) | 618 (84.4%) | 511 (69.8%) |
| **Incentive preference** | | | | | | |
| No reward expected | 542 (50.8%) | 320 (59.0%) | 510 (94.1%) | 468 (86.3%) | 465 (85.8%) | 379 (69.9%) |
| Small non-financial token appreciation | 396 (37.1%) | 213 (53.8%) | 363 (91.7%) | 331 (83.6%) | 332 (83.8%) | 267 (67.4%) |
| Financial/substantial incentive preferred | 129 (12.1%) | 44 (34.1%) | 107 (82.9%) | 89 (69.0%) | 85 (65.9%) | 65 (50.4%) |
| **Healthcare access** | | | | | | |
| **Daytime travel time to nearest healthcare facility** | | | | | | |
| Less than 30 minutes | 644 (60.4%) | 361 (56.1%) | 593 (92.1%) | 528 (82.0%) | 534 (82.9%) | 441 (68.5%) |
| Over 30 minutes | 415 (38.9%) | 214 (51.6%) | 379 (91.3%) | 352 (84.8%) | 342 (82.4%) | 268 (64.6%) |
| Prefer not to say/don't know | 8 (0.7%) | 2 (25.0%) | 8 (100.0%) | 8 (100.0%) | 6 (75.0%) | 2 (25.0%) |
| **Night-time travel time to nearest healthcare facility** | | | | | | |
| Less than 30 minutes | 611 (57.3%) | 354 (57.9%) | 566 (92.6%) | 506 (82.8%) | 509 (83.3%) | 436 (71.4%) |
| Over 30 minutes | 451 (42.3%) | 221 (49.0%) | 409 (90.7%) | 377 (83.6%) | 369 (81.8%) | 273 (60.5%) |
| Prefer not to say/don't know | 5 (0.5%) | 2 (40.0%) | 5 (100.0%) | 5 (100.0%) | 4 (80.0%) | 2 (40.0%) |
| **Digital-response readiness** | | | | | | |
| Low digital response readiness | 383 (35.9%) | 151 (39.4%) | 338 (88.3%) | 291 (76.0%) | 291 (76.0%) | 215 (56.1%) |
| High digital response readiness | 684 (64.1%) | 426 (62.3%) | 642 (93.9%) | 597 (87.3%) | 591 (86.4%) | 496 (72.5%) |
| **Donation concern types** | | | | | | |
| No concern | 461 (43.2%) | 302 (65.5%) | 446 (96.7%) | 416 (90.2%) | 432 (93.7%) | 349 (75.7%) |
| Fear or physical discomfort concern | 230 (21.6%) | 102 (44.3%) | 204 (88.7%) | 181 (78.7%) | 165 (71.7%) | 133 (57.8%) |
| Health or safety concern | 285 (26.7%) | 130 (45.6%) | 249 (87.4%) | 223 (78.2%) | 203 (71.2%) | 174 (61.1%) |
| Knowledge or eligibility concern | 280 (26.2%) | 134 (47.9%) | 255 (91.1%) | 225 (80.4%) | 217 (77.5%) | 168 (60.0%) |
| Religious or cultural concern | 33 (3.1%) | 17 (51.5%) | 21 (63.6%) | 24 (72.7%) | 20 (60.6%) | 20 (60.6%) |
| Other concern | 101 (9.5%) | 48 (47.5%) | 85 (84.2%) | 80 (79.2%) | 78 (77.2%) | 67 (66.3%) |
| **Practical barriers** | | | | | | |
| **Work schedule barrier** | | | | | | |
| No work schedule barrier | 264 (24.7%) | 194 (73.5%) | 246 (93.2%) | 239 (90.5%) | 237 (89.8%) | 231 (87.5%) |
| Work schedule barrier | 803 (75.3%) | 383 (47.7%) | 734 (91.4%) | 649 (80.8%) | 645 (80.3%) | 480 (59.8%) |
| **School schedule barrier** | | | | | | |
| No school schedule barrier | 316 (29.6%) | 210 (66.5%) | 289 (91.5%) | 280 (88.6%) | 278 (88.0%) | 247 (78.2%) |
| School schedule barrier | 751 (70.4%) | 367 (48.9%) | 691 (92.0%) | 608 (81.0%) | 604 (80.4%) | 464 (61.8%) |
| **Transport barrier** | | | | | | |
| No transport barrier | 192 (18.0%) | 130 (67.7%) | 173 (90.1%) | 170 (88.5%) | 163 (84.9%) | 150 (78.1%) |
| Transport barrier | 875 (82.0%) | 447 (51.1%) | 807 (92.2%) | 718 (82.1%) | 719 (82.2%) | 561 (64.1%) |
| **Family/childcare barrier** | | | | | | |
| No family/childcare barrier | 476 (44.6%) | 293 (61.6%) | 447 (93.9%) | 417 (87.6%) | 398 (83.6%) | 357 (75.0%) |
| Family/childcare barrier | 591 (55.4%) | 284 (48.1%) | 533 (90.2%) | 471 (79.7%) | 484 (81.9%) | 354 (59.9%) |

| Variable | Subgroup size, n (% of total sample) | High mobilisability, n (% within subgroup) | High future donation willingness, n (% within subgroup) | High trusted app installation willingness, n (% within subgroup) | High trusted-request response willingness, n (% within subgroup) | High practical flexibility, n (% within subgroup) |
|---|---|---|---|---|---|---|
| **Weather barrier** | | | | | | |
| No weather barrier | 300 (28.1%) | 204 (68.0%) | 281 (93.7%) | 266 (88.7%) | 269 (89.7%) | 237 (79.0%) |
| Weather barrier | 767 (71.9%) | 373 (48.6%) | 699 (91.1%) | 622 (81.1%) | 613 (79.9%) | 474 (61.8%) |
| **Trust in blood-request sources** | | | | | | |
| **Formal health-system trust** | | | | | | |
| Low formal health-system trust | 95 (8.9%) | 20 (21.1%) | 67 (70.5%) | 57 (60.0%) | 56 (58.9%) | 44 (46.3%) |
| High formal health-system trust | 972 (91.1%) | 557 (57.3%) | 913 (93.9%) | 831 (85.5%) | 826 (85.0%) | 667 (68.6%) |
| **Interpersonal network trust** | | | | | | |
| Low trust in interpersonal network sources | 70 (6.6%) | 23 (32.9%) | 55 (78.6%) | 44 (62.9%) | 41 (58.6%) | 34 (48.6%) |
| High interpersonal network trust | 997 (93.4%) | 554 (55.6%) | 925 (92.8%) | 844 (84.7%) | 841 (84.4%) | 677 (67.9%) |
| **Community or organisational trust** | | | | | | |
| Low trust in community/organisational sources | 170 (15.9%) | 52 (30.6%) | 143 (84.1%) | 109 (64.1%) | 109 (64.1%) | 81 (47.6%) |
| High community or organisational trust | 897 (84.1%) | 525 (58.5%) | 837 (93.3%) | 779 (86.8%) | 773 (86.2%) | 630 (70.2%) |
| **Unknown individual trust** | | | | | | |
| low trust in unknown individuals | 588 (55.1%) | 268 (45.6%) | 523 (88.9%) | 447 (76.0%) | 455 (77.4%) | 360 (61.2%) |
| High unknown individual trust | 479 (44.9%) | 309 (64.5%) | 457 (95.4%) | 441 (92.1%) | 427 (89.1%) | 351 (73.3%) |
| **Recipient-related donation willingness** | | | | | | |
| **Socially-connected recipient willingness** | | | | | | |
| Low willingness for socially-connected recipients | 30 (2.8%) | 0 (0.0%) | 9 (30.0%) | 9 (30.0%) | 4 (13.3%) | 7 (23.3%) |
| High socially-connected recipient willingness | 1,037 (97.2%) | 577 (55.6%) | 971 (93.6%) | 879 (84.8%) | 878 (84.7%) | 704 (67.9%) |
| **Anonymous stranger willingness** | | | | | | |
| Low willingness for anonymous stranger | 320 (30.0%) | 116 (36.2%) | 269 (84.1%) | 222 (69.4%) | 207 (64.7%) | 168 (52.5%) |
| High anonymous stranger willingness | 747 (70.0%) | 461 (61.7%) | 711 (95.2%) | 666 (89.2%) | 675 (90.4%) | 543 (72.7%) |
| **Emotionally prompted stranger willingness** | | | | | | |
| Low willingness for emotionally prompted stranger | 174 (16.3%) | 49 (28.2%) | 132 (75.9%) | 106 (60.9%) | 89 (51.1%) | 82 (47.1%) |
| High emotionally prompted stranger willingness | 893 (83.7%) | 528 (59.1%) | 848 (95.0%) | 782 (87.6%) | 793 (88.8%) | 629 (70.4%) |

#### Appendix 4: Regional distribution of the survey sample

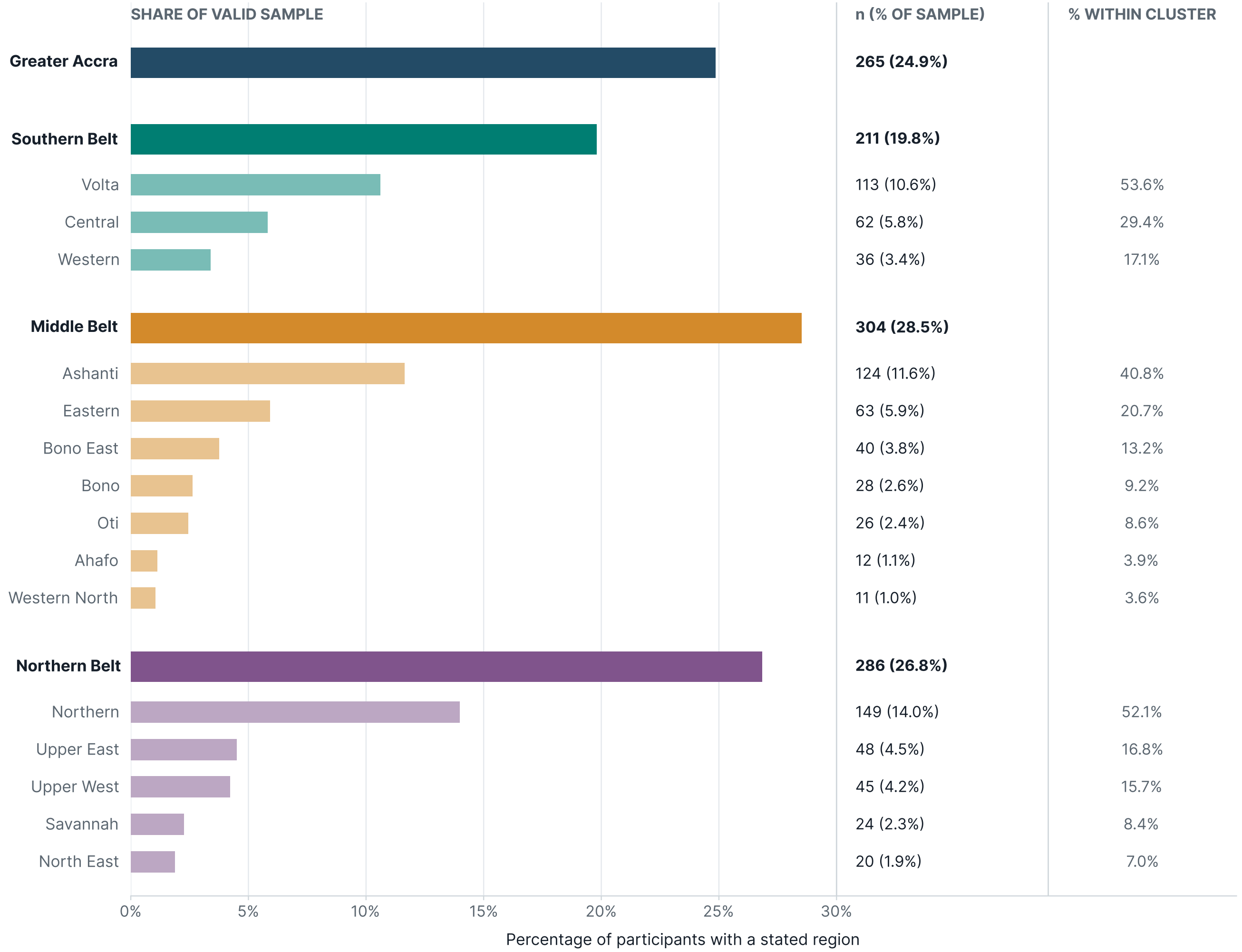

**Figure A1. Regional distribution of survey participants across Ghana.** Participants with a stated region (n=1,066) were distributed across all 16 administrative regions of Ghana. For analysis, regions were grouped into four regional clusters: Greater Accra, Southern (Coastal) Belt, Middle Belt and Northern Belt, with Greater Accra treated as a separate cluster. Bars and values in the middle column show the number and percentage of the valid sample from each region or cluster; the right-hand column shows each administrative region's percentage within its corresponding cluster. One participant who selected "Prefer not to say" for region was excluded.

#### Appendix 5: Primary bivariable screening

| Table A3. Bivariable associations with high digital urgent blood-donor mobilisability and screening of predictor variables for multivariable analysis. | | | | | | |
|---|---|---|---|---|---|---|
| **Variable** | **Subgroup n (%)** | **High mobilisability n/N (%)** | **Crude OR (95% CI)** | **Bivariable Wald p** | **Bivariable overall LRT p** | **Selected (LRT p < 0.20)** |
| **Sociodemographic characteristics** | | | | | | |
| **Gender** | | | | | | |
| Male (reference) | 715 (67.2%) | 434/715 (60.7%) | 1.00 | | **<0.001** | **Yes** |
| Female | 349 (32.8%) | 142/349 (40.7%) | 0.44 (0.34 to 0.58) | <0.001 | | |
| **Age group** | | | | | | |
| Younger (≤35) (reference) | 780 (73.1%) | 404/780 (51.8%) | 1.00 | | **0.013** | **Yes** |
| Older (>35) | 287 (26.9%) | 173/287 (60.3%) | 1.41 (1.07 to 1.86) | 0.014 | | |
| **Religion group** | | | | | | |
| Non-Christian (reference) | 276 (26.1%) | 168/276 (60.9%) | 1.00 | | **0.010** | **Yes** |
| Christian | 782 (73.9%) | 406/782 (51.9%) | 0.69 (0.52 to 0.92) | 0.010 | | |
| **Ethnicity group** | | | | | | |
| Akan (reference) | 358 (34.1%) | 176/358 (49.2%) | 1.00 | | **0.002** | **Yes** |
| Ewe | 211 (20.1%) | 106/211 (50.2%) | 1.04 (0.74 to 1.47) | 0.804 | | |
| Mole-Dagbani | 165 (15.7%) | 108/165 (65.5%) | 1.96 (1.34 to 2.87) | <0.001 | | |
| Other | 316 (30.1%) | 179/316 (56.6%) | 1.35 (1.00 to 1.83) | 0.052 | | |
| **Region cluster** | | | | | | |
| Greater Accra (reference) | 265 (24.9%) | 127/265 (47.9%) | 1.00 | | **<0.001** | **Yes** |
| Southern Belt | 211 (19.8%) | 103/211 (48.8%) | 1.04 (0.72 to 1.49) | 0.847 | | |
| Middle Belt | 304 (28.5%) | 166/304 (54.6%) | 1.31 (0.94 to 1.82) | 0.112 | | |
| Northern Belt | 286 (26.8%) | 181/286 (63.3%) | 1.87 (1.33 to 2.63) | <0.001 | | |
| **Residence type** | | | | | | |
| Urban (reference) | 412 (39.1%) | 231/412 (56.1%) | 1.00 | | 0.312 | No |
| Non-Urban | 641 (60.9%) | 339/641 (52.9%) | 0.88 (0.69 to 1.13) | 0.312 | | |
| **Education group** | | | | | | |
| Non-tertiary (reference) | 216 (20.3%) | 120/216 (55.6%) | 1.00 | | 0.618 | No |
| Tertiary | 846 (79.7%) | 454/846 (53.7%) | 0.93 (0.69 to 1.25) | 0.619 | | |
| **Economic activity status** | | | | | | |
| Inactive (reference) | 428 (40.9%) | 228/428 (53.3%) | 1.00 | | 0.670 | No |
| Active | 619 (59.1%) | 338/619 (54.6%) | 1.06 (0.82 to 1.35) | 0.670 | | |
| **Blood-donation experience, knowledge, and incentive preferences** | | | | | | |
| **Previous blood donation status** | | | | | | |
| Non-donor (reference) | 448 (42.0%) | 192/448 (42.9%) | 1.00 | | **<0.001** | **Yes** |
| Donor | 619 (58.0%) | 385/619 (62.2%) | 2.19 (1.71 to 2.81) | <0.001 | | |
| **Blood-donation knowledge** | | | | | | |
| Low (reference) | 709 (66.4%) | 360/709 (50.8%) | 1.00 | | **0.002** | **Yes** |
| High | 358 (33.6%) | 217/358 (60.6%) | 1.49 (1.15 to 1.93) | 0.002 | | |
| **Blood-donation information exposure** | | | | | | |
| Low exposure (reference) | 898 (84.2%) | 477/898 (53.1%) | 1.00 | | 0.146 | Yes |
| High exposure | 169 (15.8%) | 100/169 (59.2%) | 1.28 (0.92 to 1.79) | 0.148 | | |
| **Known transfusion experience** | | | | | | |
| No/Not Sure (reference) | 335 (31.4%) | 152/335 (45.4%) | 1.00 | | **<0.001** | **Yes** |
| Yes | 732 (68.6%) | 425/732 (58.1%) | 1.67 (1.28 to 2.16) | <0.001 | | |
| **Incentive preference** | | | | | | |
| No reward expected (reference) | 542 (50.8%) | 320/542 (59.0%) | 1.00 | | **<0.001** | **Yes** |
| Small non-financial token appreciation | 396 (37.1%) | 213/396 (53.8%) | 0.81 (0.62 to 1.05) | 0.109 | | |
| Financial/substantial incentive preferred | 129 (12.1%) | 44/129 (34.1%) | 0.36 (0.24 to 0.54) | <0.001 | | |

| | | | | | | |
|---|---|---|---|---|---|---|
| **Healthcare access** | | | | | | |
| **Daytime travel time to nearest healthcare facility** | | | | | | |
| Less than 30 minutes (reference) | 644 (60.8%) | 361/644 (56.1%) | 1.00 | | 0.152 | Yes |
| Over 30 minutes | 415 (39.2%) | 214/415 (51.6%) | 0.83 (0.65 to 1.07) | 0.152 | | |
| **Night-time travel time to nearest healthcare facility** | | | | | | |
| Less than 30 minutes (reference) | 611 (57.5%) | 354/611 (57.9%) | 1.00 | | **0.004** | **Yes** |
| Over 30 minutes | 451 (42.5%) | 221/451 (49.0%) | 0.70 (0.55 to 0.89) | 0.004 | | |
| **Digital-response readiness** | | | | | | |
| Low digital response readiness (reference) | 383 (35.9%) | 151/383 (39.4%) | 1.00 | | **<0.001** | **Yes** |
| High digital response readiness | 684 (64.1%) | 426/684 (62.3%) | 2.54 (1.96 to 3.28) | <0.001 | | |
| **Practical barriers** | | | | | | |
| **Work schedule barrier** | | | | | | |
| No work schedule barrier (reference) | 264 (24.7%) | 194/264 (73.5%) | 1.00 | | **<0.001** | **Yes** |
| Work schedule barrier | 803 (75.3%) | 383/803 (47.7%) | 0.33 (0.24 to 0.45) | <0.001 | | |
| **School schedule barrier** | | | | | | |
| No school schedule barrier (reference) | 316 (29.6%) | 210/316 (66.5%) | 1.00 | | **<0.001** | **Yes** |
| School schedule barrier | 751 (70.4%) | 367/751 (48.9%) | 0.48 (0.37 to 0.63) | <0.001 | | |
| **Transport barrier** | | | | | | |
| No transport barrier (reference) | 192 (18.0%) | 130/192 (67.7%) | 1.00 | | **<0.001** | **Yes** |
| Transport barrier | 875 (82.0%) | 447/875 (51.1%) | 0.50 (0.36 to 0.69) | <0.001 | | |
| **Family/childcare barrier** | | | | | | |
| No family/childcare barrier (reference) | 476 (44.6%) | 293/476 (61.6%) | 1.00 | | **<0.001** | **Yes** |
| Family/childcare barrier | 591 (55.4%) | 284/591 (48.1%) | 0.58 (0.45 to 0.74) | <0.001 | | |
| **Weather barrier** | | | | | | |
| No weather barrier (reference) | 300 (28.1%) | 204/300 (68.0%) | 1.00 | | **<0.001** | **Yes** |
| Weather barrier | 767 (71.9%) | 373/767 (48.6%) | 0.45 (0.34 to 0.59) | <0.001 | | |
| **Trust in blood-request sources** | | | | | | |
| **Formal health-system trust** | | | | | | |
| no formal health-system trust (reference) | 95 (8.9%) | 20/95 (21.1%) | 1.00 | | **<0.001** | **Yes** |
| Formal health-system trust | 972 (91.1%) | 557/972 (57.3%) | 5.03 (3.02 to 8.38) | <0.001 | | |
| **Interpersonal network trust** | | | | | | |
| no high trust in interpersonal network sources (reference) | 70 (6.6%) | 23/70 (32.9%) | 1.00 | | **<0.001** | **Yes** |
| Interpersonal network trust | 997 (93.4%) | 554/997 (55.6%) | 2.56 (1.53 to 4.27) | <0.001 | | |
| **Community or organisational trust** | | | | | | |
| no high trust in community/organisational sources (reference) | 170 (15.9%) | 52/170 (30.6%) | 1.00 | | **<0.001** | **Yes** |
| Community or organisational trust | 897 (84.1%) | 525/897 (58.5%) | 3.20 (2.25 to 4.55) | <0.001 | | |
| **Unknown individual trust** | | | | | | |
| does not trust unknown individuals (reference) | 588 (55.1%) | 268/588 (45.6%) | 1.00 | | **<0.001** | **Yes** |
| Unknown individual trust | 479 (44.9%) | 309/479 (64.5%) | 2.17 (1.69 to 2.78) | <0.001 | | |
| **Recipient-related donation willingness** | | | | | | |
| **Socially-connected recipient willingness** | | | | | | |
| Low willingness for socially-connected recipients (reference) | 30 (2.8%) | 0/30 (0.0%) | 1.00 | | **<0.001** | **Yes** |
| High socially-connected recipient willingness | 1,037 (97.2%) | 577/1,037 (55.6%) | Unstable | 0.999 | | |
| **Anonymous stranger willingness** | | | | | | |
| Low willingness for anonymous stranger (reference) | 320 (30.0%) | 116/320 (36.2%) | 1.00 | | **<0.001** | **Yes** |
| High anonymous stranger willingness | 747 (70.0%) | 461/747 (61.7%) | 2.83 (2.16 to 3.72) | <0.001 | | |
| **Emotionally prompted stranger willingness** | | | | | | |
| Low willingness for emotionally prompted stranger (reference) | 174 (16.3%) | 49/174 (28.2%) | 1.00 | | **<0.001** | **Yes** |
| High emotionally prompted stranger willingness | 893 (83.7%) | 528/893 (59.1%) | 3.69 (2.58 to 5.27) | <0.001 | | |

Subgroup percentages use predictor-valid denominators. Red subgroup rows indicate bivariable Wald p<0.05. Overall likelihood-ratio test (LRT) p and selection status are variable-level results.

#### Appendix 6: Primary multivariable logistic regression model diagnostics

Complete-case N=1,002; events=548 (54.7%). AUC=0.785; Hosmer-Lemeshow test p=0.239; max VIF=2.29.
Socially-connected recipient willingness was excluded from the adjusted primary model the low-willingness group contained no events. All respondents with high mobilisability also reported a high willingness to donate to socially-connected recipient.

**Table A4. Complete-case sample composition and outcome distribution for the primary multivariable logistic regression model.**

| Complete-case check | n | % | Denominator |
|---|---|---|---|
| Candidate sample available for model | 1067 | 100.0 | Candidate sample |
| Complete cases included in model | 1002 | 93.9 | Candidate sample |
| Excluded because outcome was missing | 0 | 0.0 | Candidate sample |
| Excluded because at least one model predictor was missing | 65 | 6.1 | Candidate sample |
| High-mobilisability events among complete cases | 548 | 54.7 | Complete-case sample |
| Low-mobilisability non-events among complete cases | 454 | 45.3 | Complete-case sample |

#### Appendix 7: Secondary multivariable logistic model (Component-specific exploratory analysis)

The composite mobilisability outcome was decomposed into its four original binary components. Each model used the same adjusted predictor set as the primary model, so the estimates show whether associations were concentrated in a specific operational component.

**Table A5. Model diagnostics for the four component-specific multivariable logistic regression models.**

| Component outcome | Complete-case N | Events n (%) | AUC | Hosmer-Lemeshow p | Max VIF |
|---|---|---|---|---|---|
| Future donation willingness | 1002 | 922 (92.0%) | 0.816 | 0.799 | 2.29 |
| Trusted app installation willingness | 1002 | 836 (83.4%) | 0.798 | 0.795 | 2.29 |
| Trusted-request response willingness | 1002 | 832 (83.0%) | 0.827 | 0.011 | 2.29 |
| Practical flexibility | 1002 | 671 (67.0%) | 0.772 | 0.334 | 2.29 |

**Table A6. Secondary regression models for the four original mobilisability components**

| Variable | Future donation willingness | | Trusted-app installation willingness | | Trusted-request response willingness | | Practical flexibility | |
|---|---|---|---|---|---|---|---|---|
| | AOR (95% CI) | Wald p | AOR (95% CI) | Wald p | AOR (95% CI) | Wald p | AOR (95% CI) | Wald p |
| **Sociodemographic characteristics** | | | | | | | | |
| Female (ref: Male) | **0.48 (0.28 to 0.85)** | **0.011** | **0.53 (0.35 to 0.80)** | **0.003** | 0.80 (0.52 to 1.22) | 0.298 | **0.65 (0.47 to 0.91)** | **0.011** |
| Older (>35) (ref: Younger (≤35)) | **0.48 (0.27 to 0.86)** | **0.013** | 1.14 (0.71 to 1.83) | 0.581 | 0.98 (0.60 to 1.58) | 0.928 | **1.64 (1.13 to 2.38)** | **0.010** |
| Christian (ref: Non-Christian) | 1.38 (0.65 to 2.93) | 0.398 | 1.64 (0.94 to 2.88) | 0.081 | 1.34 (0.74 to 2.40) | 0.332 | 1.11 (0.71 to 1.74) | 0.639 |
| **Ethnicity group (ref: Akan)** | | | | | | | | |
| Ewe | 0.87 (0.43 to 1.76) | 0.704 | 1.02 (0.60 to 1.74) | 0.928 | **1.86 (1.04 to 3.30)** | **0.035** | 0.91 (0.59 to 1.41) | 0.680 |
| Mole-Dagbani | 1.16 (0.39 to 3.43) | 0.787 | **2.67 (1.17 to 6.10)** | **0.020** | 1.18 (0.54 to 2.58) | 0.673 | 1.70 (0.91 to 3.19) | 0.099 |
| Other | 1.09 (0.54 to 2.21) | 0.805 | 1.14 (0.69 to 1.91) | 0.607 | 1.69 (0.99 to 2.89) | 0.056 | 1.08 (0.72 to 1.63) | 0.708 |
| **Region cluster (ref: Greater Accra)** | | | | | | | | |
| Southern Belt | 1.21 (0.57 to 2.60) | 0.622 | 1.57 (0.88 to 2.77) | 0.124 | 1.14 (0.63 to 2.07) | 0.660 | 1.06 (0.67 to 1.69) | 0.802 |
| Middle Belt | 1.18 (0.57 to 2.44) | 0.651 | 1.63 (0.95 to 2.79) | 0.073 | 1.43 (0.82 to 2.49) | 0.202 | 1.10 (0.72 to 1.69) | 0.648 |
| Northern Belt | 1.31 (0.52 to 3.30) | 0.563 | 1.28 (0.68 to 2.41) | 0.438 | 1.61 (0.82 to 3.15) | 0.167 | 1.24 (0.75 to 2.05) | 0.398 |
| Non-urban residence (ref: Urban) | 0.61 (0.34 to 1.08) | 0.092 | 0.81 (0.54 to 1.23) | 0.329 | 0.69 (0.44 to 1.06) | 0.093 | 1.22 (0.88 to 1.69) | 0.225 |
| Tertiary education (ref: Non-tertiary) | 0.85 (0.42 to 1.72) | 0.659 | 0.61 (0.36 to 1.05) | 0.073 | 1.22 (0.73 to 2.03) | 0.442 | 1.00 (0.67 to 1.48) | 0.987 |
| Economically active (ref: Inactive) | 0.73 (0.40 to 1.31) | 0.284 | 1.11 (0.74 to 1.68) | 0.610 | 0.79 (0.51 to 1.22) | 0.291 | 0.95 (0.68 to 1.31) | 0.737 |
| **Blood-donation experience, knowledge, and incentive preferences** | | | | | | | | |
| Previous donor (ref: Non-donor) | 1.46 (0.84 to 2.55) | 0.177 | 1.31 (0.87 to 1.96) | 0.196 | **3.39 (2.21 to 5.21)**† | **<0.001** | 1.37 (1.00 to 1.89) | 0.053 |
| High blood-donation knowledge (ref: Low) | 1.27 (0.71 to 2.28) | 0.422 | 1.53 (0.98 to 2.39) | 0.059 | 0.89 (0.57 to 1.39) | 0.608 | 1.38 (0.98 to 1.94) | 0.066 |
| High information exposure (ref: Low exposure) | 0.81 (0.40 to 1.62) | 0.555 | 0.84 (0.49 to 1.44) | 0.524 | 0.95 (0.54 to 1.66) | 0.846 | 1.00 (0.65 to 1.53) | 0.985 |
| Known transfusion experience (ref: No/Not Sure) | 0.81 (0.46 to 1.45) | 0.484 | 1.37 (0.92 to 2.05) | 0.122 | 1.17 (0.76 to 1.80) | 0.483 | 1.10 (0.80 to 1.53) | 0.554 |
| **Incentive preference (ref: No reward expected)** | | | | | | | | |
| Small non-financial token appreciation | 0.77 (0.43 to 1.36) | 0.365 | 0.97 (0.63 to 1.47) | 0.871 | 1.09 (0.70 to 1.68) | 0.707 | 1.25 (0.90 to 1.73) | 0.187 |
| Financial/substantial incentive preferred | 0.59 (0.28 to 1.26) | 0.174 | 0.68 (0.39 to 1.19) | 0.177 | 0.80 (0.44 to 1.45) | 0.462 | 0.81 (0.50 to 1.32) | 0.397 |
| **Healthcare access** | | | | | | | | |
| Daytime travel time over 30 min (ref: Less than 30 min) | 1.06 (0.54 to 2.10) | 0.858 | 1.41 (0.86 to 2.34) | 0.176 | 0.90 (0.53 to 1.52) | 0.693 | 1.19 (0.80 to 1.77) | 0.379 |
| Night-time travel time over 30 min (ref: Less than 30 min) | 0.71 (0.36 to 1.40) | 0.324 | 0.89 (0.55 to 1.45) | 0.645 | 0.96 (0.57 to 1.59) | 0.862 | **0.55 (0.37 to 0.80)** | **0.002** |
| **Digital response readiness** | | | | | | | | |
| High digital response readiness (ref: Low readiness) | **1.81 (1.08 to 3.06)** | **0.026** | **1.79 (1.22 to 2.64)** | **0.003** | **1.82 (1.21 to 2.74)** | **0.004** | **1.78 (1.31 to 2.43)**† | **<0.001** |
| **Practical barriers** | | | | | | | | |
| Work schedule barrier (ref: No this barrier) | 0.82 (0.38 to 1.77) | 0.610 | 0.61 (0.34 to 1.11) | 0.104 | **0.54 (0.29 to 1.00)** | **0.048** | **0.24 (0.15 to 0.40)**† | **<0.001** |
| School schedule barrier (ref: No this barrier) | 1.17 (0.62 to 2.24) | 0.626 | 0.81 (0.49 to 1.34) | 0.419 | 0.60 (0.35 to 1.03) | 0.063 | 0.82 (0.55 to 1.22) | 0.331 |
| Transport barrier (ref: No this barrier) | 1.72 (0.80 to 3.72) | 0.165 | 0.88 (0.47 to 1.63) | 0.678 | 1.28 (0.70 to 2.34) | 0.419 | 1.13 (0.70 to 1.83) | 0.614 |
| Family/childcare barrier (ref: No this barrier) | 0.67 (0.36 to 1.25) | 0.207 | 0.74 (0.48 to 1.15) | 0.185 | **1.86 (1.19 to 2.91)** | **0.006** | 0.79 (0.56 to 1.11) | 0.182 |
| Weather barrier (ref: No this barrier) | 0.97 (0.47 to 1.99) | 0.924 | 1.10 (0.66 to 1.85) | 0.705 | **0.53 (0.30 to 0.93)** | **0.027** | 0.80 (0.54 to 1.20) | 0.284 |
| **Trust in blood-request sources** | | | | | | | | |
| High formal health-system trust (ref: Low trust) | **3.85 (2.00 to 7.41)**† | **<0.001** | **2.22 (1.25 to 3.94)** | **0.007** | **2.08 (1.13 to 3.83)** | **0.019** | 1.70 (0.99 to 2.92) | 0.055 |
| High interpersonal network trust (ref: Low trust) | 1.25 (0.50 to 3.16) | 0.636 | 0.98 (0.47 to 2.01) | 0.946 | 1.15 (0.53 to 2.51) | 0.727 | 1.07 (0.55 to 2.08) | 0.830 |

| Variable | Future donation willingness | | Trusted-app installation willingness | | Trusted-request response willingness | | Practical flexibility | |
|---|---|---|---|---|---|---|---|---|
| | AOR (95% CI) | Wald p | AOR (95% CI) | Wald p | AOR (95% CI) | Wald p | AOR (95% CI) | Wald p |
| High community or organisational trust (ref: Low trust) | 0.81 (0.40 to 1.67) | 0.573 | 1.38 (0.84 to 2.26) | 0.206 | 1.17 (0.68 to 2.01) | 0.583 | 1.39 (0.89 to 2.18) | 0.149 |
| High unknown individual trust (ref: Low trust) | 1.49 (0.80 to 2.79) | 0.213 | **2.21 (1.39 to 3.52)**† | **<0.001** | 1.09 (0.68 to 1.73) | 0.726 | 1.06 (0.76 to 1.49) | 0.736 |
| **Recipient-related donation willingness** | | | | | | | | |
| High anonymous stranger willingness (ref: Low willingness) | 1.21 (0.63 to 2.34) | 0.562 | 1.57 (0.99 to 2.49) | 0.053 | **1.82 (1.14 to 2.92)** | **0.013** | **1.62 (1.11 to 2.35)** | **0.012** |
| High emotionally prompted stranger willingness (ref: Low willingness) | **2.98 (1.49 to 5.94)** | **0.002** | **2.10 (1.26 to 3.53)** | **0.005** | **4.54 (2.67 to 7.71)**† | **<0.001** | 1.53 (0.96 to 2.42) | 0.071 |

Each component model used the same adjusted predictor set as the primary model. Bold values indicate p<0.05. Superscript † indicates p<0.001.

#### Appendix 8: Donation concerns and practical barriers distributions

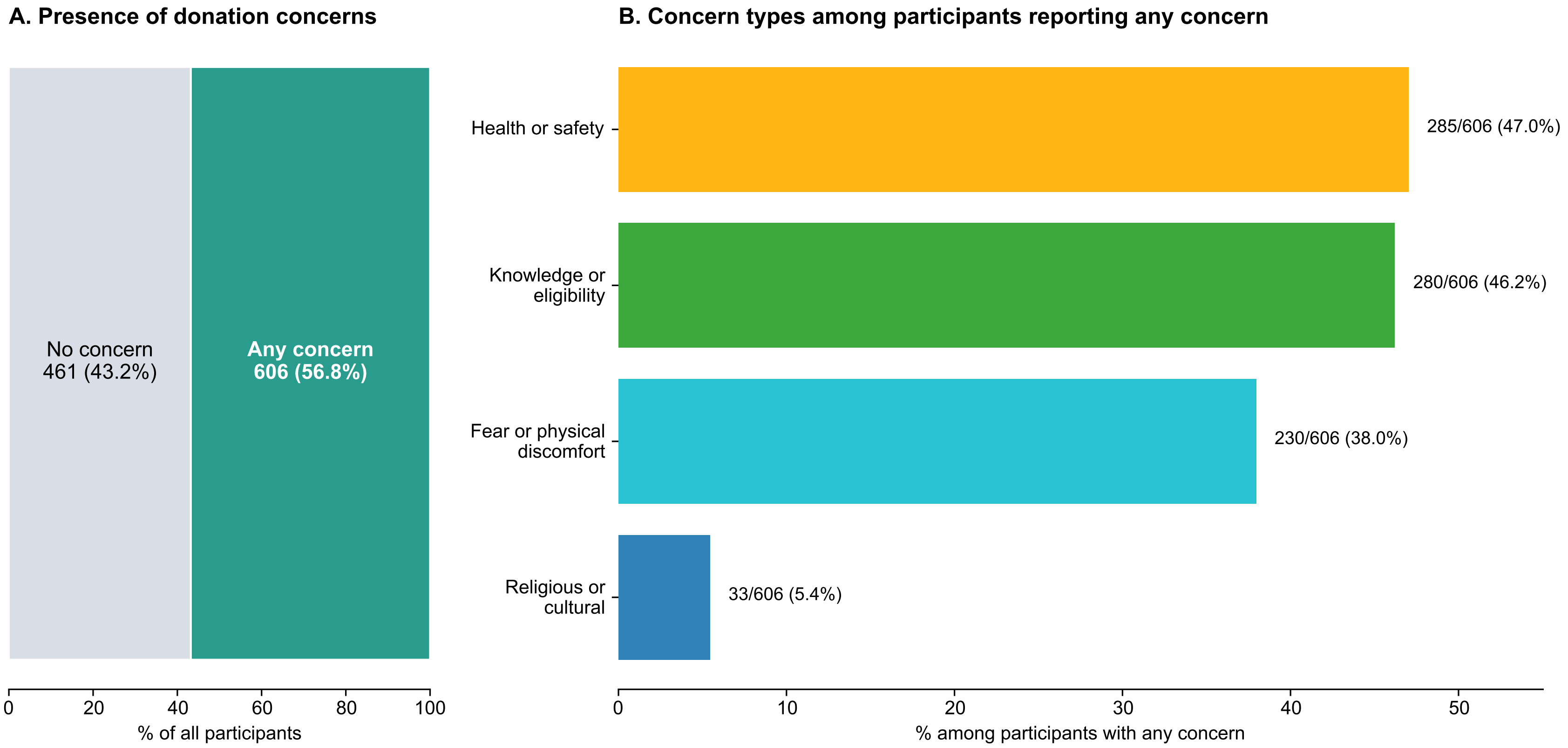

**Figure A2. Donation concerns and concern-type profile among survey participants**. (A) Distribution of participants reporting no donation concern or at least one concern in the full sample (N=1,067). (B) Number and percentage reporting each concern type among participants who reported at least one concern (n=606). Participants could report more than one concern type; therefore, percentages in panel B do not sum to 100%.

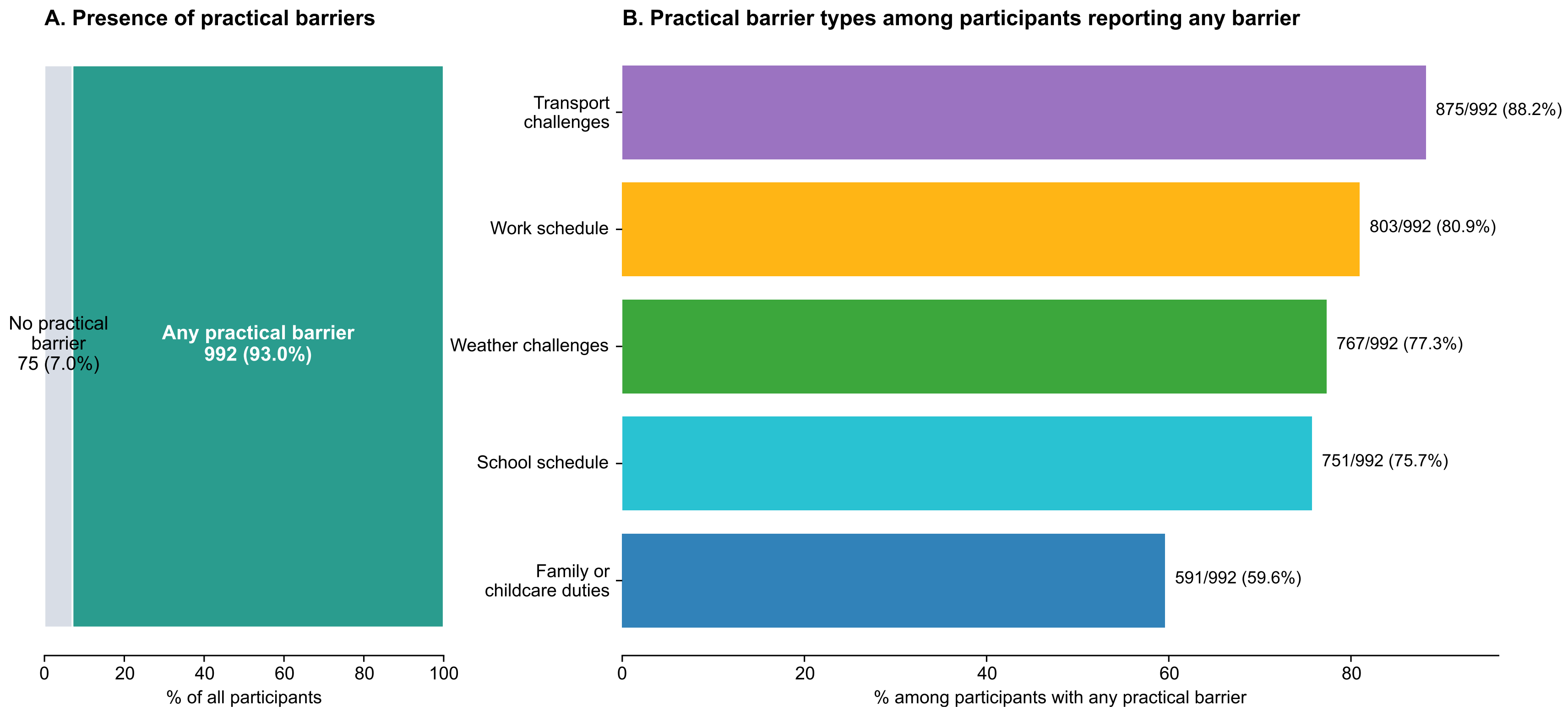

**Figure A3. Practical barriers and barrier-type profile among survey participants.** (A) Distribution of participants reporting no practical barrier or at least one practical barrier in the full sample (N=1,067). (B) Number and percentage reporting each barrier type among participants who reported at least one practical barrier (n=992). Participants could report more than one barrier type; therefore, percentages in panel B do not sum to 100%.

### Appendix 9-10: Supplementary sensitivity analyses

#### Appendix 9: Sensitivity model 1 (Conditional concern-type model)

The conditional model was restricted to respondents reporting any donation concern (N=606). Model complete-case N=564; events=263 (46.6%). AUC=0.795; Hosmer-Lemeshow test p=0.257; max VIF=2.53. Previous donor status was not included in this follow-up model by design.

**Table A7. Complete-case sample composition and outcome distribution for the conditional concern-type sensitivity model.**

| Complete-case check | n | % | Denominator |
|---|---|---|---|
| Candidate sample available for model | 606 | 100.0 | Candidate sample |
| Complete cases included in model | 564 | 93.1 | Candidate sample |
| Excluded because outcome was missing | 0 | 0.0 | Candidate sample |
| Excluded because at least one model predictor was missing | 42 | 6.9 | Candidate sample |
| High-mobilisability events among complete cases | 263 | 46.6 | Complete-case sample |
| Lower-mobilisability non-events among complete cases | 301 | 53.4 | Complete-case sample |

**Table A8. Adjusted concern-type model among respondents reporting any donation concern**

| Variable | Adjusted OR (95% CI) | Wald p |
|---|---|---|
| **Sociodemographic characteristics** | | |
| Female (ref: Male) | **0.53 (0.35 to 0.81)** | **0.004** |
| Older (>35) (ref: Younger (≤35)) | 1.27 (0.76 to 2.12) | 0.367 |
| Christian (ref: Non-Christian) | **1.83 (1.02 to 3.29)** | **0.043** |
| **Ethnicity group (ref: Akan)** | | |
| Ewe | 1.00 (0.54 to 1.84) | 0.996 |
| Mole-Dagbani | **3.11 (1.37 to 7.05)** | **0.007** |
| Other | 1.40 (0.80 to 2.46) | 0.234 |
| **Region cluster (ref: Greater Accra)** | | |
| Southern Belt | 1.05 (0.56 to 1.98) | 0.884 |
| Middle Belt | 1.09 (0.61 to 1.97) | 0.769 |
| Northern Belt | 1.13 (0.59 to 2.15) | 0.709 |
| Non-urban residence (ref: Urban) | 0.85 (0.55 to 1.30) | 0.447 |
| Tertiary education (ref: Non-tertiary) | 0.81 (0.48 to 1.36) | 0.423 |
| Economically active (ref: Inactive) | 0.90 (0.58 to 1.39) | 0.632 |
| **Blood-donation experience, knowledge, and incentive preferences** | | |
| High blood-donation knowledge (ref: Low) | 1.42 (0.87 to 2.33) | 0.164 |
| High information exposure (ref: Low exposure) | 0.78 (0.43 to 1.38) | 0.389 |
| Known transfusion experience (ref: No/Not Sure) | 1.19 (0.76 to 1.87) | 0.451 |
| **Incentive preference (ref: No reward expected)** | | |
| Small non-financial token appreciation | 1.22 (0.79 to 1.90) | 0.372 |
| Financial/substantial incentive preferred | 0.69 (0.36 to 1.34) | 0.274 |
| **Healthcare access** | | |
| Daytime travel time over 30 minutes (ref: Less than 30 minutes) | 1.12 (0.66 to 1.90) | 0.663 |

| Variable | Adjusted OR (95% CI) | Wald p |
|---|---|---|
| Night-time travel time over 30 minutes (ref: Less than 30 minutes) | 0.65 (0.39 to 1.11) | 0.116 |
| **Digital-response readiness** | | |
| High digital response readiness (ref: Low readiness) | **2.26 (1.46 to 3.48)**† | **<0.001** |
| **Donation concern types** | | |
| Knowledge or eligibility concern (ref: No this concern) | 1.20 (0.77 to 1.85) | 0.420 |
| Fear or physical discomfort concern (ref: No this concern) | 0.96 (0.63 to 1.46) | 0.838 |
| Religious or cultural concern (ref: No this concern) | **3.61 (1.29 to 10.14)** | **0.015** |
| Practical inconvenience concern (ref: No this concern) | 1.11 (0.66 to 1.86) | 0.692 |
| Health or safety concern (ref: No this concern) | 1.28 (0.84 to 1.93) | 0.248 |
| **Practical barriers** | | |
| Work schedule barrier (ref: No this barrier) | **0.45 (0.26 to 0.78)** | **0.005** |
| School schedule barrier (ref: No this barrier) | 0.72 (0.43 to 1.20) | 0.207 |
| Transport barrier (ref: No this barrier) | 1.14 (0.61 to 2.16) | 0.678 |
| Family/childcare barrier (ref: No this barrier) | 1.14 (0.72 to 1.81) | 0.586 |
| Weather barrier (ref: No this barrier) | 0.68 (0.40 to 1.16) | 0.158 |
| **Trust in blood-request sources** | | |
| High formal health-system trust (ref: Low trust) | **4.16 (1.93 to 8.97)**† | **<0.001** |
| High interpersonal network trust (ref: Low trust) | 0.74 (0.31 to 1.75) | 0.491 |
| High community or organisational trust (ref: Low trust) | 1.47 (0.81 to 2.65) | 0.203 |
| High unknown individual trust (ref: Low trust) | 1.39 (0.89 to 2.18) | 0.143 |
| **Recipient-related donation willingness** | | |
| High anonymous stranger willingness (ref: Low willingness) | **2.12 (1.30 to 3.46)** | **0.003** |
| High emotionally prompted stranger willingness (ref: Low willingness) | **2.25 (1.17 to 4.35)** | **0.015** |

Bold values indicate $p<0.05$. Superscript † indicates $p<0.001$. Reference groups are shown in parentheses.

#### Appendix 10: Sensitivity model 2 (Item-level sensitivity model)

The sensitivity model replaced high digital response readiness with its three components and replaced grouped domains with item-level indicators. Complete-case N=1,002; events=548; AUC=0.801; Hosmer-Lemeshow test p=0.916; max VIF=2.30.

**Table A9. Complete-case sample composition and outcome distribution for the item-level sensitivity model.**

| Complete-case check | n | % | Denominator |
|---|---|---|---|
| Candidate sample available for model | 1067 | 100.0 | Candidate sample |
| Complete cases included in model | 1002 | 93.9 | Candidate sample |
| Excluded because outcome was missing | 0 | 0.0 | Candidate sample |
| Excluded because at least one model predictor was missing | 65 | 6.1 | Candidate sample |
| High-mobilisability events among complete cases | 548 | 54.7 | Complete-case sample |
| Lower-mobilisability non-events among complete cases | 454 | 45.3 | Complete-case sample |

**Table A10. Item-level sensitivity model with item-level readiness, trust, and recipient-related willingness predictors**

| Variable | Adjusted OR (95% CI) | Wald p |
|---|---|---|
| **Sociodemographic characteristics** | | |
| Female (ref: Male) | **0.60 (0.43 to 0.84)** | **0.003** |
| Older (>35) (ref: Younger (≤35)) | 1.26 (0.88 to 1.81) | 0.211 |
| Christian (ref: Non-Christian) | 1.32 (0.85 to 2.03) | 0.216 |
| **Ethnicity group (ref: Akan)** | | |
| Ewe | 0.99 (0.64 to 1.53) | 0.951 |
| Mole-Dagbani | 1.64 (0.89 to 3.02) | 0.112 |
| Other | 1.21 (0.80 to 1.83) | 0.365 |
| **Region cluster (ref: Greater Accra)** | | |
| Southern Belt | 1.18 (0.73 to 1.90) | 0.501 |
| Middle Belt | 1.23 (0.80 to 1.89) | 0.341 |
| Northern Belt | 1.40 (0.86 to 2.29) | 0.179 |
| Non-urban residence (ref: Urban) | 0.90 (0.65 to 1.24) | 0.530 |
| Tertiary education (ref: Non-tertiary) | 0.79 (0.53 to 1.17) | 0.240 |
| Economically active (ref: Inactive) | 0.88 (0.64 to 1.22) | 0.449 |
| **Blood-donation experience, knowledge, and incentive preferences** | | |
| Previous donor (ref: Non-donor) | 1.34 (0.97 to 1.86) | 0.074 |
| High blood-donation knowledge (ref: Low) | **1.49 (1.06 to 2.08)** | **0.020** |
| High information exposure (ref: Low exposure) | 1.00 (0.66 to 1.53) | 0.992 |
| Known transfusion experience (ref: No/Not Sure) | 1.21 (0.87 to 1.69) | 0.250 |
| **Incentive preference (ref: No reward expected)** | | |
| Small non-financial token appreciation | 1.05 (0.76 to 1.45) | 0.761 |
| Financial/substantial incentive preferred | 0.68 (0.40 to 1.14) | 0.143 |
| **Healthcare access** | | |
| Daytime travel time over 30 minutes (ref: Less than 30 minutes) | 0.96 (0.65 to 1.42) | 0.835 |
| Night-time travel time over 30 minutes (ref: Less than 30 minutes) | 0.69 (0.47 to 1.02) | 0.060 |
| **Digital-response readiness components** | | |
| Can install/use new apps alone or with minimal help (ref: needs more than minimal help) | 0.88 (0.36 to 2.14) | 0.781 |
| Likely to answer trusted-source call (ref: not likely to answer trusted-source call) | **2.65 (1.64 to 4.30)**[†] | **<0.001** |
| Checks important notifications within 30 seconds (ref: checks important notifications after 30 seconds or misses them) | **1.83 (1.29 to 2.58)**[†] | **<0.001** |
| **Practical barriers** | | |

| Variable | Adjusted OR (95% CI) | Wald p |
|---|---|---|
| Work schedule barrier (ref: No this barrier) | **0.43 (0.28 to 0.65)**† | **<0.001** |
| School schedule barrier (ref: No this barrier) | 0.73 (0.50 to 1.07) | 0.103 |
| Transport barrier (ref: No this barrier) | 0.90 (0.57 to 1.43) | 0.654 |
| Family/childcare barrier (ref: No this barrier) | 1.04 (0.74 to 1.45) | 0.837 |
| Weather barrier (ref: No this barrier) | 0.80 (0.55 to 1.17) | 0.253 |
| **Trust in blood-request sources (item-level indicators)** | | |
| High trust in hospitals/clinics (ref: Low trust) | **3.26 (1.69 to 6.30)**† | **<0.001** |
| High trust in family/friends (ref: Low trust) | 0.78 (0.41 to 1.49) | 0.452 |
| High trust in unknown individuals (ref: Low trust) | 1.24 (0.87 to 1.76) | 0.228 |
| High trust in ordinary community members (ref: Low trust) | 0.90 (0.59 to 1.39) | 0.640 |
| High trust in blood-donation groups (ref: Low trust) | 1.26 (0.89 to 1.79) | 0.192 |
| High trust in religious groups (ref: Low trust) | 1.15 (0.70 to 1.88) | 0.587 |
| High trust in community leaders/local officials (ref: Low trust) | 0.93 (0.57 to 1.53) | 0.779 |
| **Recipient-related donation willingness (item-level indicators)** | | |
| Willingness for family/friends (ref: low willingness) | **5.72 (1.16 to 28.25)** | **0.032** |
| Willingness for admired public figures/celebrities (ref: low willingness) | **1.73 (1.14 to 2.62)** | **0.010** |
| Willingness for community members (ref: low willingness) | **1.84 (1.05 to 3.24)** | **0.034** |
| Willingness for anonymous stranger (ref: low willingness) | 1.24 (0.83 to 1.84) | 0.288 |
| Willingness for emotionally prompted stranger (ref: low willingness) | 1.46 (0.86 to 2.45) | 0.158 |

Bold values indicate p<0.05. Superscript † indicates p<0.001. Reference groups are shown in parentheses.

#### Appendix 11: Comparison of the survey sample with national benchmarks (Ghana 2021 census data)

Comparisons are intended primarily to provide context on the composition of the survey sample. Two-sided one-sample exact binomial tests were used to compare each survey proportion with the corresponding national benchmark. Because the survey used non-probability online recruitment and was not designed to generate nationally representative population estimates, p values should be interpreted as indicating differences from the benchmark within this sample rather than as a formal assessment of population representativeness. Survey denominators vary by characteristic because “Prefer not to say” and “I don’t know” responses were treated as missing. National benchmark denominators vary according to the population covered by the relevant Ghana 2021 Population and Housing Census table. Adult-specific benchmarks were used where available or derivable; religion, ethnicity, region and residence benchmarks were based on the populations reported in the corresponding census tables. The economic-activity benchmark relates to people aged ≥15 years.

**Table A11. Comparison of survey participant characteristics with national benchmarks from the 2021 Ghana Population and Housing Census.**

| Characteristic | Survey respondents, n/N (%) | Ghana national benchmark, n/N (%) | Difference, percentage points | p value |
|---|---|---|---|---|
| **Sex** | | | | |
| Male | 715/1,064 (67.2%) | 8,664,693/17,931,673 (48.3%) | +18.9 | **< 0.001** |
| Female | 349/1,064 (32.8%) | 9,266,980/17,931,673 (51.7%) | -18.9 | **< 0.001** |
| **Age** | | | | |
| 18–35 years | 780/1,067 (73.1%) | 9,772,721/17,931,673 (54.5%) | +18.6 | **< 0.001** |
| >35 years | 287/1,067 (26.9%) | 8,158,952/17,931,673 (45.5%) | -18.6 | **< 0.001** |
| **Religion** | | | | |
| Christian | 782/1,058 (73.9%) | 21,932,708/30,753,327 (71.3%) | +2.6 | 0.066 |
| Non-Christian | 276/1,058 (26.1%) | 8,820,619/30,753,327 (28.7%) | -2.6 | 0.066 |
| **Ethnicity (Top 3)** | | | | |
| Akan | 358/1,050 (34.1%) | 13,925,576/30,484,536 (45.7%) | -11.6 | **< 0.001** |
| Ewe | 211/1,050 (20.1%) | 3,902,009/30,484,536 (12.8%) | +7.3 | **< 0.001** |
| Mole-Dagbani | 165/1,050 (15.7%) | 5,638,881/30,484,536 (18.5%) | -2.8 | **0.021** |
| **Region** | | | | |
| Greater Accra | 265/1,066 (24.9%) | 5,455,692/30,832,019 (17.7%) | +7.2 | **< 0.001** |
| All other regions | 801/1,066 (75.1%) | 25,376,327/30,832,019 (82.3%) | -7.2 | **< 0.001** |
| **Residence** | | | | |
| Urban | 412/1,053 (39.1%) | 17,472,530/30,832,019 (56.7%) | -17.5 | **< 0.001** |
| Non-urban (survey) / rural (census) | 641/1,053 (60.9%) | 13,359,489/30,832,019 (43.3%) | +17.5 | **< 0.001** |
| **Education** | | | | |
| Tertiary | 846/1,062 (79.7%) | 2,288,434/17,931,673 (12.8%) | +66.9 | **< 0.001** |
| Non-tertiary | 216/1,062 (20.3%) | 15,643,239/17,931,673 (87.2%) | -66.9 | **< 0.001** |
| **Economic activity** | | | | |
| Active | 619/1,047 (59.1%) | 11,541,355/19,873,607 (58.1%) | +1.0 | 0.511 |
| Inactive | 428/1,047 (40.9%) | 8,332,252/19,873,607 (41.9%) | -1.0 | 0.511 |

Difference is the survey percentage minus the benchmark percentage. p values are from two-sided, unadjusted one-sample exact binomial tests comparing each survey proportion with the corresponding national benchmark proportion. Bold indicates $p < 0.05$; values below 0.001 are shown as <0.001. As the survey is a non-probability sample, p values should be interpreted descriptively.

#### Appendix 12: STROBE Statement—Checklist of items that should be included in reports of *cross-sectional studies*

| | Item No | Recommendation | Section/paragraph |
|---|---|---|---|
| **Title and abstract** | 1 | (*a*) Indicate the study's design with a commonly used term in the title or the abstract | Title; Abstract |
| | | (*b*) Provide in the abstract an informative and balanced summary of what was done and what was found | Abstract |
| **Introduction** | | | |
| Background/rationale | 2 | Explain the scientific background and rationale for the investigation being reported | Introduction (paragraphs 1-6) |
| Objectives | 3 | State specific objectives, including any prespecified hypotheses | Introduction (paragraph 7) |
| **Methods** | | | |
| Study design | 4 | Present key elements of study design early in the paper | Methods (Study setting and design, paragraphs 1-3) |
| Setting | 5 | Describe the setting, locations, and relevant dates, including periods of recruitment, exposure, follow-up, and data collection | Methods (Study setting and design, paragraphs 1-3) |
| Participants | 6 | (*a*) Give the eligibility criteria, and the sources and methods of selection of participants | Methods (Study setting and design, paragraphs 1 and 3) |
| Variables | 7 | Clearly define all outcomes, exposures, predictors, potential confounders, and effect modifiers. Give diagnostic criteria, if applicable | Methods (Outcome definition, paragraph 1; Explanatory variables, paragraphs 1–5) |
| Data sources/ measurement | 8* | For each variable of interest, give sources of data and details of methods of assessment (measurement). Describe comparability of assessment methods if there is more than one group | Methods (Study setting and design, paragraph 2; Outcome definition, paragraph 1; Explanatory variables, paragraphs 1-5) |
| Bias | 9 | Describe any efforts to address potential sources of bias | Methods (Study setting and design, paragraphs 1-3); Discussion (Strengths and limitations, paragraphs 2-3) |
| Study size | 10 | Explain how the study size was arrived at | Methods (Study setting and design, paragraph 1) |
| Quantitative variables | 11 | Explain how quantitative variables were handled in the analyses. If applicable, describe which groupings were chosen and why | Methods (Outcome definition, paragraph 1; Explanatory variables, paragraphs 1-5) |
| Statistical methods | 12 | (*a*) Describe all statistical methods, including those used to control for confounding | Methods (Statistical analysis, paragraphs 1-2) |
| | | (*b*) Describe any methods used to examine subgroups and interactions | Methods (Statistical analysis, paragraph 1-2) |
| | | (*c*) Explain how missing data were addressed | Methods (Statistical analysis, paragraph 1) |
| | | (*d*) If applicable, describe analytical methods taking account of sampling strategy | Methods (Study setting and design, paragraph 1; Statistical analysis, paragraph 1) |
| | | (*e*) Describe any sensitivity analyses | Methods (Statistical analysis, paragraph 2); Results (Factors associated with high digital urgent blood-donor mobilisability, paragraph 4; Donation concerns and practical barriers across selected participant groups, paragraph 4) |
| **Results** | | | |

| | | | |
|---|---|---|---|
| Participants | 13* | (a) Report numbers of individuals at each stage of study—eg numbers potentially eligible, examined for eligibility, confirmed eligible, included in the study, completing follow-up, and analysed | Results (Participant characteristics and distribution of mobilisation outcomes, paragraph 1) |
| | | (b) Give reasons for non-participation at each stage | Methods (Study setting and design, paragraph 3) |
| | | (c) Consider use of a flow diagram | n/a |
| Descriptive data | 14* | (a) Give characteristics of study participants (eg demographic, clinical, social) and information on exposures and potential confounders | Results (Participant characteristics and distribution of mobilisation outcomes, paragraph 1; Table 1; Table 2) |
| | | (b) Indicate number of participants with missing data for each variable of interest | Results (Table 1; Factors associated with high digital urgent blood-donor mobilisability, paragraph 1) |
| Outcome data | 15* | Report numbers of outcome events or summary measures | Results (Participant characteristics and distribution of mobilisation outcomes, paragraph 1; Figure 1; Table 1) |
| Main results | 16 | (*a*) Give unadjusted estimates and, if applicable, confounder-adjusted estimates and their precision (eg, 95% confidence interval). Make clear which confounders were adjusted for and why they were included | Methods (Statistical analysis, paragraph 1); Results (Factors associated with high digital urgent blood-donor mobilisability, paragraphs 1-5; Table 2) |
| | | (*b*) Report category boundaries when continuous variables were categorized | Methods (Outcome definition and Explanatory variables); Results (Tables 1-2) |
| | | (*c*) If relevant, consider translating estimates of relative risk into absolute risk for a meaningful time period | n/a |
| Other analyses | 17 | Report other analyses done—eg analyses of subgroups and interactions, and sensitivity analyses | Results (Component-specific associations across the mobilisation pathway, paragraphs 1-2; Donation concerns and practical barriers across selected participant groups, paragraphs 1-4; Figures 2-3) |
| **Discussion** | | | |
| Key results | 18 | Summarise key results with reference to study objectives | Discussion (paragraph 1); Conclusion |
| Limitations | 19 | Discuss limitations of the study, taking into account sources of potential bias or imprecision. Discuss both direction and magnitude of any potential bias | Discussion (Strengths and limitations, paragraphs 2-3) |
| Interpretation | 20 | Give a cautious overall interpretation of results considering objectives, limitations, multiplicity of analyses, results from similar studies, and other relevant evidence | Discussion (paragraphs 2–8; Strengths and limitations, paragraph 3); Conclusion |
| Generalisability | 21 | Discuss the generalisability (external validity) of the study results | Discussion (Strengths and limitations, paragraphs 2-3) |
| **Other information** | | | |
| Funding | 22 | Give the source of funding and the role of the funders for the present study and, if applicable, for the original study on which the present article is based | Funding statement in the end matter |

*Give information separately for exposed and unexposed groups.
